# Network meta-analysis made simple: a composite likelihood approach

**DOI:** 10.1101/2024.06.19.24309163

**Authors:** Yu-Lun Liu, Bingyu Zhang, Haitao Chu, Yong Chen

## Abstract

Network meta-analysis, also known as mixed treatments comparison meta-analysis or multiple treatments meta-analysis, extends conventional pairwise meta-analysis by simultaneously synthesizing multiple interventions in a single integrated analysis. Despite the growing popularity of network meta-analysis within comparative effectiveness research, it comes with potential challenges. For example, within-study correlations among treatment comparisons are rarely reported in the published literature. Yet, these correlations are pivotal for valid statistical inference. As demonstrated in earlier studies, ignoring these correlations can inflate mean squared errors of the resulting point estimates and lead to inaccurate standard error estimates. This paper introduces a composite likelihood-based approach that ensures accurate statistical inference without requiring knowledge of the within-study correlations. The proposed method is computationally robust and efficient, with substantially reduced computational time compared to the state-of-the-science methods implemented in R packages. The proposed method was evaluated through extensive simulations and applied to two important applications including a network meta-analysis comparing interventions for primary open-angle glaucoma, and another comparing treatments for chronic prostatitis and chronic pelvic pain syndrome.

**Highlights:** *What is already known?:*

- Network meta-analysis extends conventional pairwise meta-analysis by simultaneously synthesizing multiple interventions in a single integrated analysis.
- A significant challenge in network meta-analysis is the lack of reported within-study correlations among treatment comparisons in the published studies.

*What is new?:*

- We propose a new method for network meta-analysis that ensures vaild statistical inference without the need for knowledge of within-study correlations.
- The proposed method employs a composite likelihood and a sandwich-type robust variance estimator, offering a computationally efficient and scalable solution, particularly for network meta-analysis with a large number of treatments and studies.

*Potential impact for *Research Synthesis Methods* readers:*

- The proposed method can be easily applied to any univariate network meta-analysis project without requiring knowledge of within-study correlations among treatment comparisons.

## 1 Introduction

Meta-analysis is a widely used tool in systematic reviews for combining and contrasting multiple studies to obtain overall estimates of the relative effects in the target population.^1, 2^ The methodology of the pairwise meta-analysis, which only focuses on comparing an intervention with a reference (e.g., control or placebo), has been well developed.^3, 4^ For many medical conditions, there are often more than two interventions of interest. In such situations, performing isolated pairwise meta-analysis might not adequately represent the comprehensive landscape of interventions nor guide the selection of optimal treatment to maximize patient benefits.^5, 6^ Furthermore, it is often unrealistic to anticipate that there is at least a head-to-head trial comparing any two interventions of interest.

Network meta-analysis (NMA), coined by Lumley back in 2002,^7^ also known as multiple treatments meta-analysis or mixed treatments comparison, is a state-of-the-science technique for making inferences with multiple treatments. This approach enables the comparison of diverse treatment subsets across various trials. A notable application of NMA is the comparison of treatment options for depression conducted by Cipriani and his colleagues,^8^ which provided a comprehensive summary of the relative efficacy and safety of 21 antidepressant drugs based on all available studies to date. Their insights have the potential to influence clinical practices, impacting on millions of individuals who suffer from depression globally. Essentially, the rationale of NMA is to expand pairwise meta-analysis to simultaneously compare multiple treatments and produce consistent estimates of relative treatment effects by synthesizing both direct and indirect clinical evidence in a single integrated analysis.^6^

Over the last two decades, the literature has seen a plethora of methodological developments in NMAs, mostly focusing on meta-regression and Bayesian hierarchical models, among others.^9–12^ For instance, the estimation of indirect evidence in NMAs was first derived through a meta-regression model,^7, 13^ wherein various treatment comparisons were treated as covariates in a model. Yet, such an approach can be challenging to estimate between-study variance, especially for sparse networks, so that the between-study heterogeneity variance is often assumed to be common across all treatment comparisons in a network.^7, 13, 14^ On the other hand, the assumption of common between-study heterogeneity variance can lead to inaccurate estimates.^15^ A shift towards the Bayesian hierarchical model, championed by Lu and his colleagues,^16, 17^ has received significant interest. Their subsequent work^18^ further discussed how the consistency equations imposed restrictions on between-study heterogeneity of each treatment comparison and used the spherical parameterization based on Cholesky decomposition to implement the constraints.

In addition to the contrast-based NMA (CB-NMA), which focuses on the (weighted) average of study-specific relative effects by assuming fixed study-specific intercepts, the arm-based NMA (AB-NMA) models study-specific absolute effects and assumes random intercepts. This offers greater flexibility in estimands, including both the population-averaged absolute and relative effects.^19–23^ Alternatively, NMAs can be conceptualized as a multivariate meta-analysis.^24^ Other existing models for implementing NMAs include electrical networks and graph-theoretical methods^25, 26^ under fixed-effects or random-effects models. In addition to the classical framework of synthesizing point estimates, a novel confidence distribution framework based on a sample-dependent distribution function has been proposed.^27^

Despite the popularity of NMAs, an important challenge has not been fully addressed, which is the unknown or unreported within-study correlations. Specifically, estimates of contrasts between each pair of treatment comparisons are correlated within a multi-arm study when these contrasts involve a common comparator. On the other hand, within-study correlations are rarely reported in the published trials.^28^ When conducting an NMA, ignoring within-study correlations can lead to biased estimates of relative treatment effects; particularly, these estimates were also found to have increased mean-square errors and standard errors.^28, 29^

When the impact of within-study correlations is non-ignorable, several methods have been proposed and used to obtain estimates of within-study correlations. First, the availability of individual participant-level data allows to compute the within-study correlations directly between treatment comparisons in each trial;^30^ however, it is uncommon in meta-analysis to have the individual participant-level data for all trials. Study investigators are often unable to provide information about within-study correlations even if we make requests directly.^31^ Second, an alternative method, known as the Pearson correlation method, proposed by Kirkham et al.,^32^ can be implemented in the formulation of multivariate meta-analysis. Third, Riley et al.^33^ proposed a single correlation parameter to capture both within-study and between-study correlations in the setting of multivariate meta-analysis. As pointed out by Riley et al.,^28^ the impact of within-study correlations is relative to the magnitude of between-study variation. In other words, when total variation in estimated effect sizes across studies, as the sum of within-study and between-study covariance, is dominated by within-study variation, the impact of within-study correlations can be substantial and ignoring the unknown within-study correlations can lead to misleading results.^33^ Other methods such as Bayesian approaches^34^ have also been proposed.

Even though the within-study correlations could be estimated by the abovementioned methods, each of them requires additional assumptions and constraints along with high computational complexity to ensure that estimations of within-study variance-covariance matrices are valid and positive definite, particularly for Bayesian methods using Markov chain Monte Carlo algorithms. One of the existing methods to resolve such an issue is to restrict the range of correlation coefficients in each study from a truncated prior distribution so that the positive definiteness of the variance-covariance matrix is guaranteed.^35^ Other alternatives, such as Cholesky parameterization and spherical decomposition,^36^ have been employed to ensure positive-definite variance-covariance matrices for meta-analysis and network meta-analysis under a Bayesian framework. These methods, however, might be more difficult to implement in NMAs as the number of studies and treatments in a network grows.

To overcome the aforementioned challenges, we propose a new method without imposing any additional assumptions beyond those in a standard NMA. Compared to the conventional approach implementing NMAs with the standard full likelihood, our proposed method does not require knowledge of the typically unreported within-study correlations among treatment comparisons. Using composite likelihood^37, 38^ and the finite-sample corrected variance estimator,^39, 40^ our proposed method can lead to valid estimated effect sizes with coverage probabilities close to the nominal level. We also derived the corresponding algorithm which is computationally efficient and scalable to a large number of treatments in NMAs. Unlike the state-of-the-art methods whose computational time increases exponentially with respect to the number of treatments and studies, the computational time of our algorithm increases linearly with respect to the number of treatments and is nearly invariant with respect to the number of studies. Further, our algorithm avoids the issue of singular covariance estimates, which is a known practical issue for conducting multivariate or network meta-analysis.^29, 31, 41–44^

The rest of the paper is organized as follows. In Section 2 we give an overview of the two motivating examples, namely, a network meta-analysis comparing interventions for primary open-angle glaucoma, and a network meta-analysis comparing treatments for chronic prostatitis and chronic pelvic pain syndrome. In Section 3 we formulate the proposed method and introduce a treatment ranking procedure, while in Section 4 we describe a series of simulation studies to illustrate the concerns of conventional NMA models, and to examine the statistical properties of the proposed method. In Section 5 we present the applications of the proposed method to the two motivating examples. We conclude with a discussion and key messages in Section 6.

## 2 Two Motivating Examples

### 2.1 Comparison of interventions for primary open-angle glaucoma

Li et al.^45^ and Wang et al.^46^ conducted a network meta-analysis to compare all first-line treatments against primary open-angle glaucoma or ocular hypertension. Glaucoma is a disease of the optic nerve characterized by optic nerve head changes and associated visual field defects.^47^ This NMA consisted of 125 trials comparing 14 active drugs and a placebo in subjects with primary open-angle glaucoma or ocular hypertension. The studies were collected from 1983 to 2016 through the Cochran Register of Controlled Trials, Drugs@FDA, and ClinicalTrials.gov;^46^ a total of 22,656 participants were included in these studies. Figure 1(a) visualizes the data structure. Specifically, the 14 active drugs were divided into four major drug classes, including *α*-2 adrenergic agonist, *β*-blocker, carbonic anhydrase inhibitor, and prostaglandin analog. This network consisted of 114 two-arm studies, 10 three-arm studies, and 1 four-arm study. The primary outcome of interest was the difference in mean increased intraocular pressure (IOP) measured by any method at 3 months in continuous millimeters of mercury unit. The original NMA analysis^45^ employed a Bayesian hierarchical model with the Markov chain Monte Carlo technique.^16, 17^ Their analysis focused on modeling the between-study variance-covariance matrix, assuming either a homogeneous or heterogeneous structure, rather than the within-study variance-covariance matrix. A ranking of treatments was produced through the surface under the cumulative ranking curve.^48^ The study found all active drugs were clinically effective compared with placebo in reducing IOP at 3 months, with bimatoprost, latanoprost, and travoprost ranked as the first, second, and third most efficacious drugs, respectively, in lowering IOP at 3 months.

**Figure 1:**
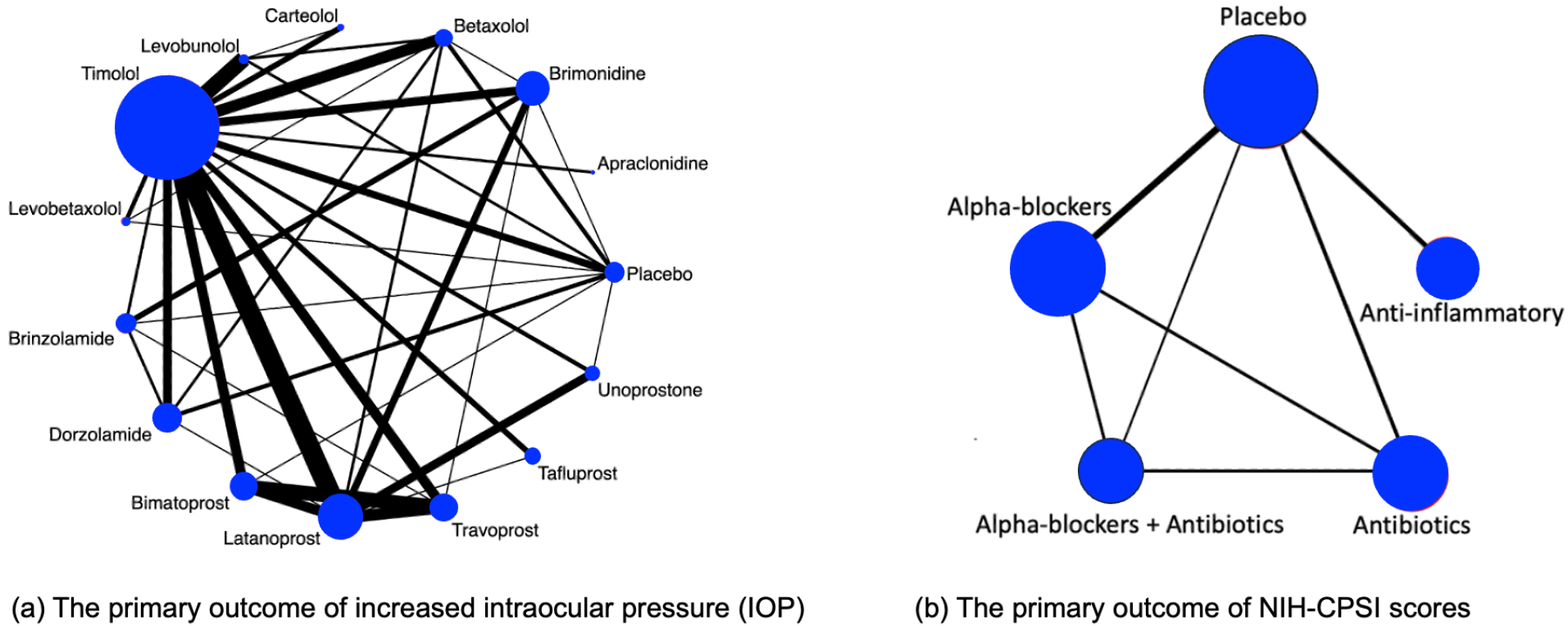
Illustration of evidence network diagrams. The size of each node is proportional to the number of participants assigned to each treatment. Solid lines represent direct comparisons between treatments in trials, with line thickness proportional to the number of trials directly comparing each pair of treatments.

As described in the Introduction section, an important limitation of this motivating example is the unknown or unreported within-study correlations. Specifically, contrast treatment estimates for the IOP outcome may be potentially correlated within a trial; for example, the two drugs, bimatoprost and travoprost, were both against latanoprost within the same trial in five studies.^49–53^ These within-study correlations among treatment comparisons were not reported, and individual participant-level data were unavailable for computing such correlations.

### 2.2 Comparison of treatments for the chronic prostatitis and chronic pelvic pain syndrome

Thakkinstian et al.^54^ performed a network meta-analysis to determine the effectiveness of multiple pharmacological therapies in improving chronic prostatitis symptoms in patients with chronic prostatitis and chronic pelvic pain syndrome (CP/CPPS). CP/CPPS is a common disorder characterized by two major clinical manifestations, including pelvic pain and lower urinary tract symptoms.^55^ The identified studies were collected from the Medicine and EMBASE databases up to 13 January 2011, and enrolled a total of 1,669 participants.

Figure 1(b) visualizes the data structure. The primary outcome of interest was the symptom score measured by the National Institutes of Health Chronic Prostatitis Symptom Index (NIH-CPSI), especially consisting of total symptoms, pain, voiding, and quality of life scores.^56^ The dataset consisted of 19 published trials comparing five treatment regimens: including placebo, any *α*-blockers (terazosin, doxazosin, tamsulosin, alfuzosin, silodosin), any antibiotics (ciprofloxacin, levofloxacin, tetracycline), anti-inflammatory/immune modulatory agents (steroidal and non-steroidal anti-inflammatory drugs, glycosaminoglycans, phytotherapy, and tanezumab), and a combination of antibiotics and *α*-blockers. Treatment comparisons among the five treatments were conducted by Thakkinstian et al.^54^ using an NMA approach; their findings suggested that *α*-blockers, antibiotics, and/or anti-inflammatory/immune modulatory agents were more efficacious in improving the total NIH-CPSI symptom scores compared to placebo. In this example, within-study correlations among treatment comparisons were not available for any of the included studies.

## 3 Methods

In this section, we introduce the proposed composite likelihood-based method for conducting network meta-analysis without the need for knowledge of within-study correlations. Throughout this paper, we focus on the contrast-based model,^13^ although our method can be extended to arm-based models.^11, 21–23^ The contrast-based model employs a two-stage estimation approach; in the first stage, the estimated effects comparing all possible intervention options in studies are computed, along with their associated standard errors from the contrast-level data, and in the second stage, the effect estimates are analyzed using a normal approximation likelihood.

### 3.1 Notations and model specification

Suppose that a network consists of *m* studies (*i* = 1, … , *m*) comparing a set of treatment options 𝒦 = *{*0, 1, 2, … , *K}* of (*K* + 1) treatments. Each design (*d* = 1, … , *D*) corresponds to a subset of 𝒦_*d*_ treatments, i.e., 𝒦_*d*_ ∈ 𝒦, and let *m*_*d*_ be the number of studies in design *d*. Under the assumptions of consistency, a network with (*K* + 1) treatments contains *K* basic parameters. These parameters are frequently taken to be the relative effects of each treatment versus a reference (or common comparator). In this paper, we do not assume that all studies utilize the same reference treatment. Instead, our proposed method incorporates all observed treatment comparisons, ensuring that reported treatment effects and standard errors contribute to the estimation of objective parameters in the proposed composite likelihood function. Moreover, the estimation does not depend on the choice of a common reference.

Suppose 𝒯 = *{jj*′*}*_*j*,*j*_′_*∈K*,*j′≠j*_ = *{*01, 02, … , (*K−*1)*K}* is the set of *N* observed treatment comparisons. Let *y*_*i*,*jj*_′ be an observed treatment effect comparing treatment *j* to treatment *j*′ in the *i*th study. Let 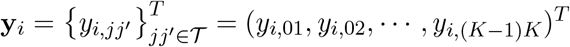 be a vector of the observed contrast of treatments, along with a vector of the associated standard errors 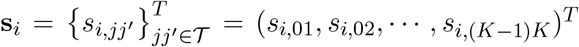. The observed relative treatment effects in an NMA are modeled via a random-effects framework,

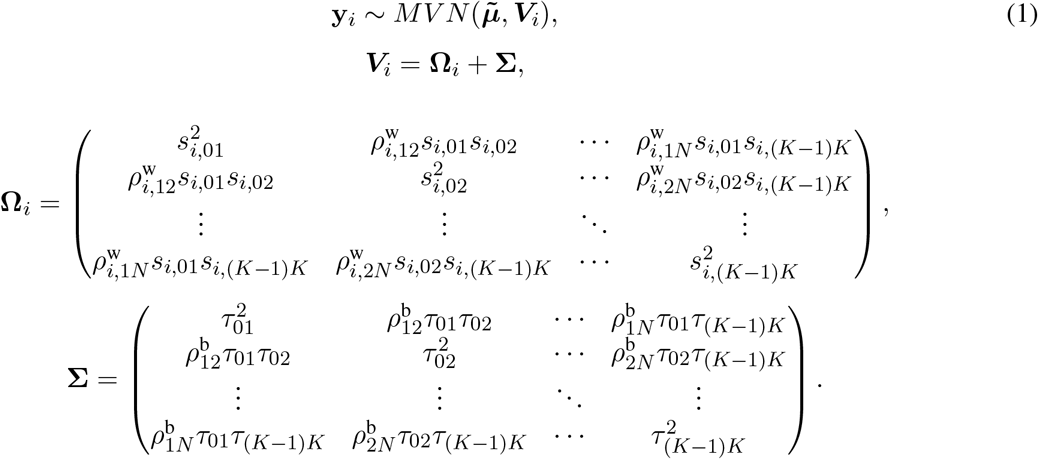

Here, 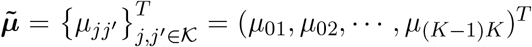 a vector of true population relative treatment effect sizes. The ***V***_*i*_ matrix indicates that the total variability affecting summary measures in each study is the sum of both within-study and between-study variance-covariance matrices. For the within-study variance-covariance 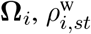 (with 1 ≤ *s < t* ≤ *N* ) refers to the within-study correlation, which is rarely reported in published literature or even calculated in each study. We assume that 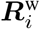 is the within-study correlation matrix, where the diagonal elements are equal to 1 and the off-diagonal elements are 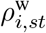 for 1 ≤ *s < t* ≤ *N* . For the between-study variance-covariance matrix **Σ**, 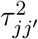represents the heterogeneity variance of the outcome comparing treatment *j* and treatment *j*′, and 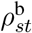 denotes the between-study correlation for 1 ≤ *s < t* ≤ *N* . We assume that ***R***^b^ is the between-study correlation matrix, where the diagonal elements are equal to 1 and the off-diagonal elements are 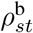 for 1 ≤ *s < t* ≤ *N* .

### 3.2 Proposed method

Let ℳ_*jj*′_ be the subset of studies that report effect sizes and standard errors of the outcome for the treatment comparison between *j* and *j*′. Let 𝓁(***η***) be the log composite likelihood function of the model defined in Equation (1) given the observed data (**y**_*i*_, **s**_*i*_). We have

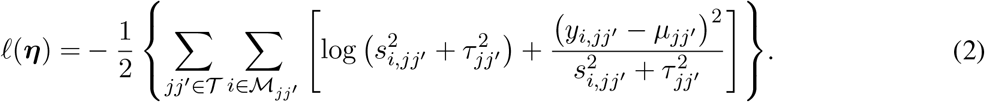

In order to fit an NMA, it is indispensable that the consistency equation is satisfied as follows, *µ*_*jj*_′ = *µ*_*jk*_ *− µ*_*j*_′_*k*_ , ∀*k≠ j, j*′. Throughout this section, we choose treatment ‘0’ as a common reference. Then, in Equation (2), we only need to estimate the parameters ***η*** = (***µ***^*T*^ , (***τ*** ^2^)^*T*^ )^*T*^ , where 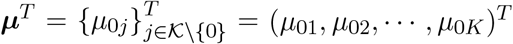 and 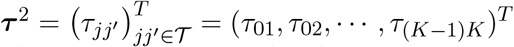 . We obtain the estimates of these parameters by maximizing the log composite likelihood function,

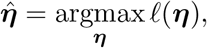

where the estimator 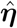 is asymptotically normal as *m* → ∞. The asymptotic variance-covariance matrix of ***η*** can be estimated by a sandwich-type estimator of form ***V*** = ***I***^*−*1^**Λ*I***^*−*1^, with 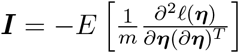 and 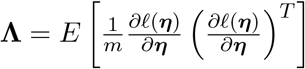. Here, we opted for a sandwich-type variance estimator, which offers two key advantages in network meta-analyses. First, it is robust to dependence. Even though the composite likelihood method assumes independence between treatment effects within a study, the sandwich estimator can partially account for this dependence. It achieves this by incorporating additional information during variance-covariance estimation, leading to more accurate standard errors for treatment effects. Second, the sandwich-type estimator is generally robust to model misspecification of covariance structures. This robustness is particularly beneficial for handling the complex data structures often encountered in network meta-analyses. However, it is important to note that the underlying marginal model itself cannot be misspecified. We also note that the restricted maximum likelihood (REML) estimator produces the same asymptotic distribution as the maximum likelihood estimator by incorporating the extra term of

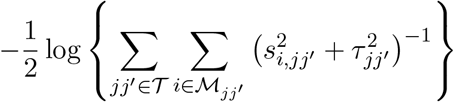

in Equation (2). Additionally, we notice that ***µ*** is information orthogonal to ***τ*** ^2^. Assuming the information matrix is 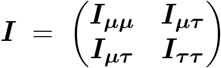 with 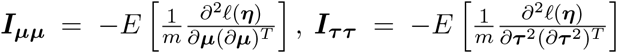 , and 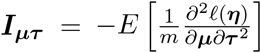. The off-diagonal element of the information matrix, ***I*** , satisfies ***I***_***µτ***_ = 0, implying that ***µ*** and ***τ*** ^2^ are information orthogonal. Under this orthogonality property, the variance-covariance matrix of 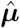 involves the information of ***µ*** alone and can be simplified as 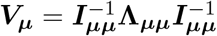 , with 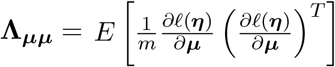. The asymptomatic variance-covariance matrix is estimated by its empirical variance-covariance matrix 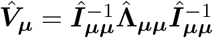 , where Î_***µµ***_ and 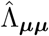 are the submatrices of ***Î*** and 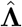 , respectively. As elucidated by Liang and Zeger,^57^ the orthogonality property implies that the between-study variance estimates have a limited impact on the estimation of effect sizes ***µ***. A detailed description of robust sandwich-type variance estimation is provided in Appendices 1 and 2 of the Supplementary Materials.

The efficient and iterative algorithm for parameter estimation can be implemented by maximizing the log composite likelihood function in Equation (2), as described in Algorithm 1. More specifically, when ***τ***^2^ is fixed at some value of 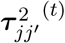 , the parameters ***µ*** can be estimated by solving a system of linear equations. In other words, maximizing 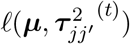 over ***µ*** yields

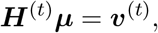

where ***µ*** is the solution of the above system of linear equations, and

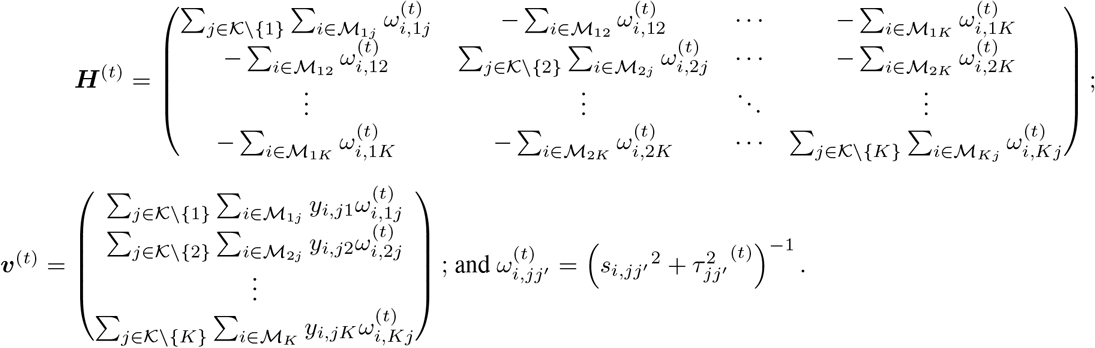

The proof of convergence for Algorithm 1 is provided in Appendix 3 of the Supplementary Materials. Additionally, a detailed description of testing for inconsistency can be found in Appendix 4 of the Supplementary Materials. The R codes for the 3-arm study is publicly available on GitHub: https://github.com/Penncil/xmeta/tree/master/R/CLNMA.equal.tau.R.

#### Algorithm 1 An efficient and simple algorithm for univariate NMA

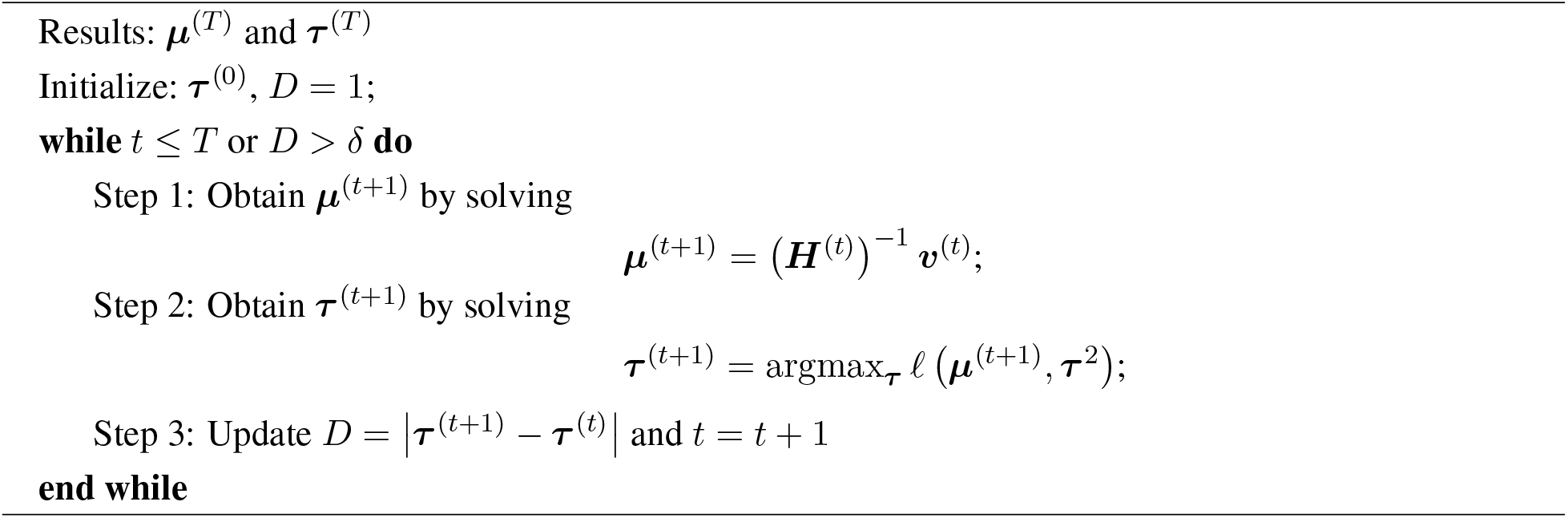

### 3.3 Treatment ranking

The hierarchy of comparable interventions can be computed by incorporating the NMA estimates obtained from the proposed composite likelihood-based method. Among various approaches to treatment ranking, the most commonly employed method relies on ranking probabilities; the probabilities for each treatment can be placed at a specific ranking position, e.g., best, second best, third best treatment, and so forth, in comparison to all other treatments in a network. These approaches for treatment ranking include the surface under the cumulative ranking curve (SUCRA)^48^ and P-score techniques,^58^ among others. In this paper, we adopted the surface under the cumulative ranking curve method.^48^ By incorporating the NMA estimates obtained from our proposed method into the SUCRA, we can properly account for the uncertainty in the estimates of relative treatment ranking. Specifically, for each treatment *j* out of the (*K* + 1) competing treatments, the SUCRA is calculated as follows:

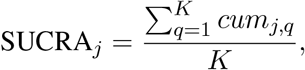

where *cum*_*j*,*q*_ refers to the cumulative probability of being among the *q* best treatment (*q* = 1, 2, … , *K* + 1). A SUCRA value of 1 indicates that the treatment is ranked as the best, while a value of 0 indicates the treatment is ranked as the worst.

## 4 Simulation Study

In this section, our objective is to assess the impact of within-study correlations on pooled estimates when employing the proposed composite likelihood-based method. We conducted extensive simulation studies to evaluate the performance of the proposed method across various scenarios, varying factors such as the within-study correlations, the between-study heterogeneity variance, and the number of studies. Furthermore, we compared the computational time of the proposed method with the existing NMA methods implemented in the R packages.

### 4.1 Data-generating mechanisms

For the simulation study, we considered a contrast-based NMA consisting of a three-arm design (i.e., *A, B*, and *C*; *A* is treated as a reference) with respect to a single continuous outcome of primary interest. The simulated data was generated via the model as the form of

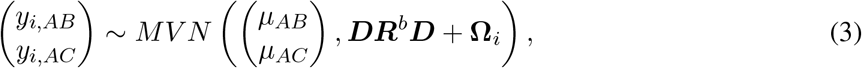

where 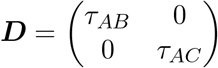 ,*R*^*b*^represents the 2 *×* 2 between-study correlation matrix, and **Ω**_*i*_ denotes the 2 *×* 2 within-study variance-covariance matrix. Within the simulated network, we assumed the consistency equation, in terms of *µ*_*BC*_ = *µ*_*AC*_ *− µ*_*AB*_. As suggested by Lu and Ades,^17^ the between-study heterogeneity variance 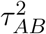 can be defined by the following relationship: 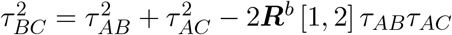 , where 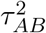 and 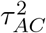 are the variances of random quantities *µ*_*AB*_ and *µ*_*AC*_, respectively. These variances are interpreted as the random effects of treatment *B* and *C* relative to a common comparator *A*.

The model parameters described above varied during simulations and were as follows. Two scenarios were considered. The first scenario used a common between-study heterogeneity variance for all treatment comparisons with *τ*_*AB*_ = *τ*_*AC*_ = 0.5, and the off-diagonal elements of ***R***^*b*^ were set to 0.1, in which ***R***^*b*^ was guaranteed to be positive semi-definite. Despite the assumption of a common between-study heterogeneity variance is widely used in practice, it remains a strong assumption. Thus, we relaxed it in the second scenario, where unequal between-study heterogeneity variance was considered with *τ*_*AB*_ = 0.7 and *τ*_*AC*_ = 0.5, respectively, and the off-diagonal elements of ***R***^*b*^ were again set to 0.1. Under both scenarios, within-study correlations were set at small (0.2) and medium (0.5) magnitudes to explore their impact. Both scenarios reflected a low-to-moderate level of heterogeneity, ranging between 20% and 35% of the total variance; in other words, between-study variance was not so large as to completely dominate the within-study variance. We generated closed loops with equal two-arm and three-arm studies into the desired network of studies, with *m*_*AB*_ = *m*_*AC*_ = *m*_*BC*_ = *m*_*ABC*_ = *m*. The true treatment effects for *AB* and *AC* comparisons were mimicked by the IOP data as described in Section 2 and set as *µ*_*AB*_ = *−*2 and *µ*_*AC*_ = *−*4, and *µ*_*BC*_ was obtained through the equation *µ*_*BC*_ = *µ*_*AB*_ *− µ*_*AC*_. The simulated sizes for studies in NMAs were set to *m* = 5, 10, 15, 20, 25, and 50. For each simulation setting, 1, 000 NMA datasets were generated. Using the model parameters described above, continuous data were generated from the multivariate normal distribution in Equation (3). The simulation study was conducted using R software, version 4.2.1.

### 4.2 Simulation results

We evaluated the performance of our proposed method by examining treatment effect estimates, in terms of bias, empirical standard error (ESE), model-based standard error (MBSE), as well as coverage of 95% confidence intervals.

Figure 2 displays the computational time for various NMA approaches. The currently available R packages include ‘*gemtc*’^59^ and ‘*netmeta*’,^60^ in which the ‘*gemtc*’ package employs the Bayesian NMA and the Markov chain Monte Carlo (MCMC), while ‘*netmeta*’ is designed based on a frequentist random-effects NMA model. We found that initial values could significantly impact the execution time of both ‘*gemtc*’ and the proposed method, whereas ‘*netmeta*’ was less affected by this issue. As the number of treatment comparisons and studies increased, differences in computational time among the three methods became more pronounced. Even though ‘*gemtc*’ and ‘*netmeta*’ could be done with minimal computational time for the scenarios with fewer studies, their computational time could increase dramatically with an increasing number of treatment comparisons and studies. Conversely, the proposed method generally yielded consistent performance in computational time, irrespective of the number of treatment comparisons or studies. Detailed computational time is summarized in Table S1 of the Supplementary Materials.

**Figure 2:**
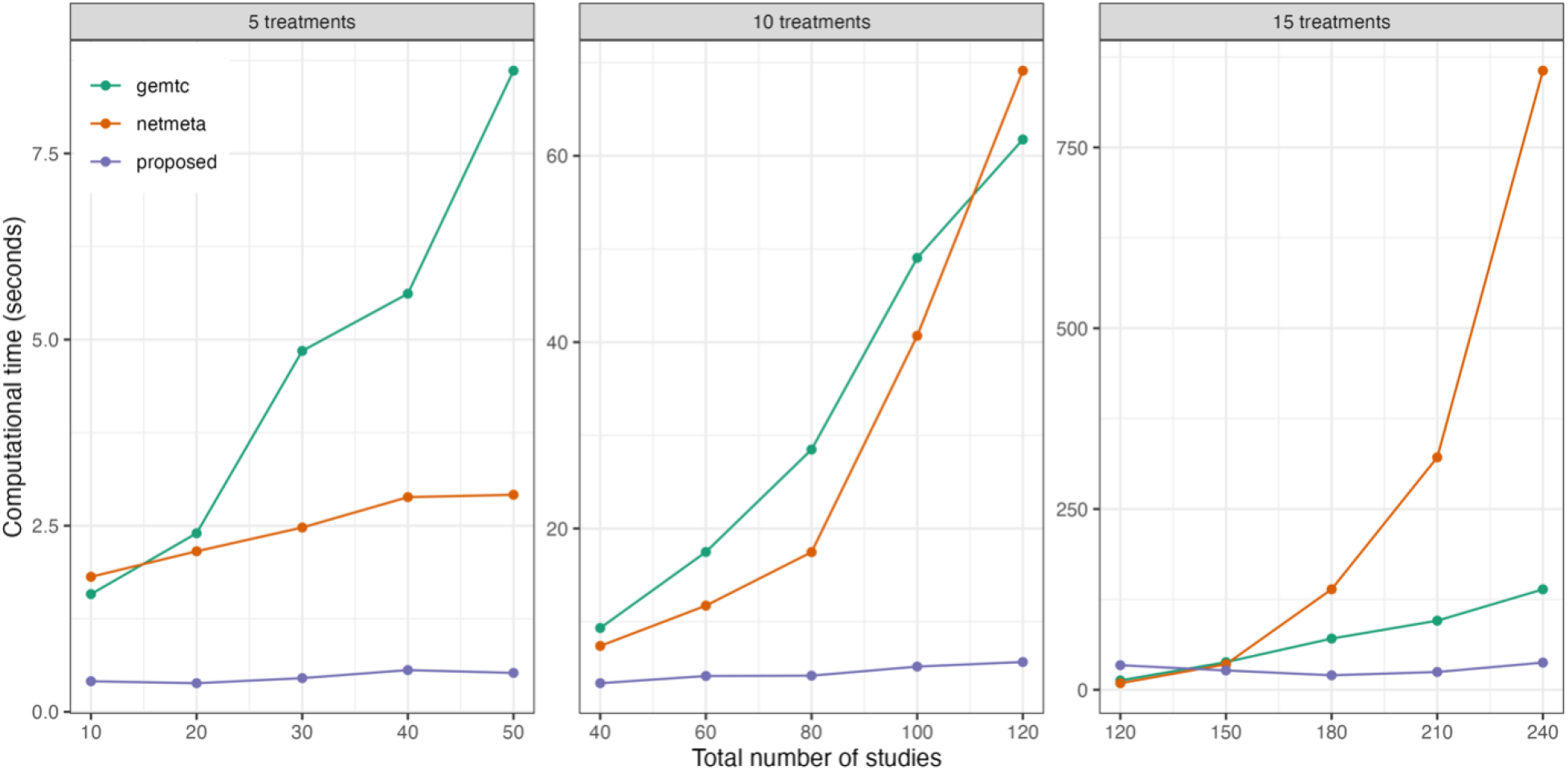
Comparisons of computational time for the proposed method and two existing methods implemented in the R packages ‘*gemtc*’ and ‘*netmeta*’, with varying numbers of treatments and studies.

The upper panel of Table 1 summarizes simulation results for treatment comparisons *AB* and *AC* in the scenario with common between-study heterogeneity. Overall, we observed that the proposed composite likelihood-based method yielded approximately unbiased pooled estimates for *AB* and *AC* treatment comparisons in most simulation settings. It did not exhibit discernible patterns in *AB* and *AC* treatment estimates across different magnitudes of within-study correlations (i.e., 0.2 and 0.5); in other words, it appeared that the magnitude of within-study correlations had a limited impact on the pooled estimates. The confidence intervals computed by the robust sandwich-type variance method demonstrated acceptable coverage probabilities ranging from 88% to 94%, relying on the number of studies. It was interesting to note that the model-based standard errors appeared somewhat smaller than their empirical standard errors. One of the possible explanations for this phenomenon was that the proposed method based on composite likelihood provided more efficient inference in large sample settings. However, as widely acknowledged in the literature,^61–66^ the variance based on the sandwich-type estimation may be underestimated when the number of studies is below 50 for continuous outcomes.

**Table 1:** Summary of 1,000 simulations with *m* = 5, 10, 15, 20, 25 and 50: bias (Bias), empirical standard error (ESE), model-based standard error (MBSE), and coverage probability (CP) of pooled estimates of *AB* and *AC* treatment comparisons. Upper panel (Scenario 1): the data-generation mechanism was through common between-study heterogeneity variance. Lower panel (Scenario 2): the data-generation mechanism was through unequal between-study heterogeneity variance. All results were based on the proposed method without any corrections.

| Within-study correlation | Number of studies | AB comparison |  |  |  | AC comparison |  |  |  |
| --- | --- | --- | --- | --- | --- | --- | --- | --- | --- |
|  |  | Bias | ESE | MBSE | CP | Bias | ESE | MBSE | CP |
| Scenario 1: $\tau_{AB} = \tau_{AC} = 0.5$, $\mu_{AB} = -2$, and $\mu_{AC} = -4$ | | | | | | | | | |
| 0.2 | 5 | 0.0118 | 0.3102 | 0.2860 | 0.908 | -0.0038 | 0.3261 | 0.2872 | 0.900 |
|  | 10 | 0.0044 | 0.2361 | 0.2074 | 0.909 | 0.0037 | 0.2468 | 0.2102 | 0.890 |
|  | 15 | -0.0002 | 0.1925 | 0.1713 | 0.905 | -0.0010 | 0.1932 | 0.1714 | 0.914 |
|  | 20 | -0.0018 | 0.1646 | 0.1484 | 0.911 | -0.0012 | 0.1647 | 0.1487 | 0.922 |
|  | 25 | 0.0008 | 0.1428 | 0.1333 | 0.937 | -0.0003 | 0.1456 | 0.1332 | 0.920 |
|  | 50 | 0.0012 | 0.1030 | 0.0945 | 0.930 | 0.0025 | 0.1059 | 0.0946 | 0.924 |
| 0.5 | 5 | -0.0079 | 0.3456 | 0.2778 | 0.876 | -0.0026 | 0.3284 | 0.2794 | 0.883 |
|  | 10 | 0.0218 | 0.2347 | 0.2010 | 0.890 | 0.0130 | 0.2279 | 0.2007 | 0.903 |
|  | 15 | 0.0013 | 0.1817 | 0.1668 | 0.923 | -0.0015 | 0.1948 | 0.1657 | 0.896 |
|  | 20 | -0.0011 | 0.1594 | 0.1445 | 0.921 | -0.0087 | 0.1572 | 0.1443 | 0.925 |
|  | 25 | -0.0033 | 0.1386 | 0.1292 | 0.939 | -0.0073 | 0.1410 | 0.1293 | 0.926 |
|  | 50 | 0.0064 | 0.1007 | 0.0917 | 0.928 | 0.0031 | 0.0998 | 0.0917 | 0.928 |
| Scenario 2: $\tau_{AB} = 0.7$, $\tau_{AC} = 0.5$, $\mu_{AB} = -2$, and $\mu_{AC} = -4$ | | | | | | | | | |
| 0.2 | 5 | -0.0100 | 0.3473 | 0.3082 | 0.892 | 0.0077 | 0.3393 | 0.2967 | 0.896 |
|  | 10 | 0.0108 | 0.2487 | 0.2223 | 0.915 | -0.0074 | 0.2396 | 0.2138 | 0.911 |
|  | 15 | -0.0003 | 0.2032 | 0.1843 | 0.925 | 0.0012 | 0.1888 | 0.1762 | 0.927 |
|  | 20 | -0.0059 | 0.1744 | 0.1589 | 0.930 | -0.0064 | 0.1691 | 0.1522 | 0.918 |
|  | 25 | -0.0019 | 0.1565 | 0.1429 | 0.917 | 0.0021 | 0.1484 | 0.1363 | 0.915 |
|  | 50 | -0.0017 | 0.1146 | 0.1017 | 0.919 | -0.0016 | 0.0996 | 0.0973 | 0.942 |
| 0.5 | 5 | 0.0057 | 0.3550 | 0.3022 | 0.891 | 0.0066 | 0.3210 | 0.2883 | 0.910 |
|  | 10 | -0.0024 | 0.2454 | 0.2175 | 0.910 | -0.0013 | 0.2265 | 0.2088 | 0.923 |
|  | 15 | 0.0081 | 0.2086 | 0.1786 | 0.898 | 0.0134 | 0.1932 | 0.1711 | 0.911 |
|  | 20 | 0.0061 | 0.1786 | 0.1545 | 0.904 | 0.0050 | 0.1660 | 0.1480 | 0.913 |
|  | 25 | 0.0001 | 0.1610 | 0.1394 | 0.904 | 0.0011 | 0.1447 | 0.1332 | 0.911 |
|  | 50 | 0.0057 | 0.1092 | 0.0987 | 0.915 | 0.0076 | 0.1012 | 0.0946 | 0.933 |

To resolve this issue, several alternative bias-corrected sandwich estimators have been proposed to improve such a small sample performance, such as KC-corrected sandwich estimator^67^ and MD-corrected sandwich estimator,^63^ among others. As indicated by Li and Redden,^66^ no single bias-corrected sandwich estimator is universally superior; however, a rule of thumb is to choose the KC-corrected method when the coefficient of variation is less than 0.6. Through simulation studies, we evaluated whether the coverage probabilities of 95% confidence intervals obtained by the proposed method were improved after corrections. Figure 3 displays comparisons of coverage probabilities using the proposed method with and without the KC-corrected and MD-corrected techniques for situations with the number of studies being 5, 10, 15, 20, 25 (or even 50). We found that the proposed method with corrections exhibited higher coverage probabilities compared to the proposed method without any corrections, particularly when the number of studies was relatively small (e.g., 5, 10, and 15 studies). Detailed results with the KC-corrected and MD-corrected methods are provided in Tables S2 and S3 of the Supplementary Materials, respectively.

**Figure 3:**
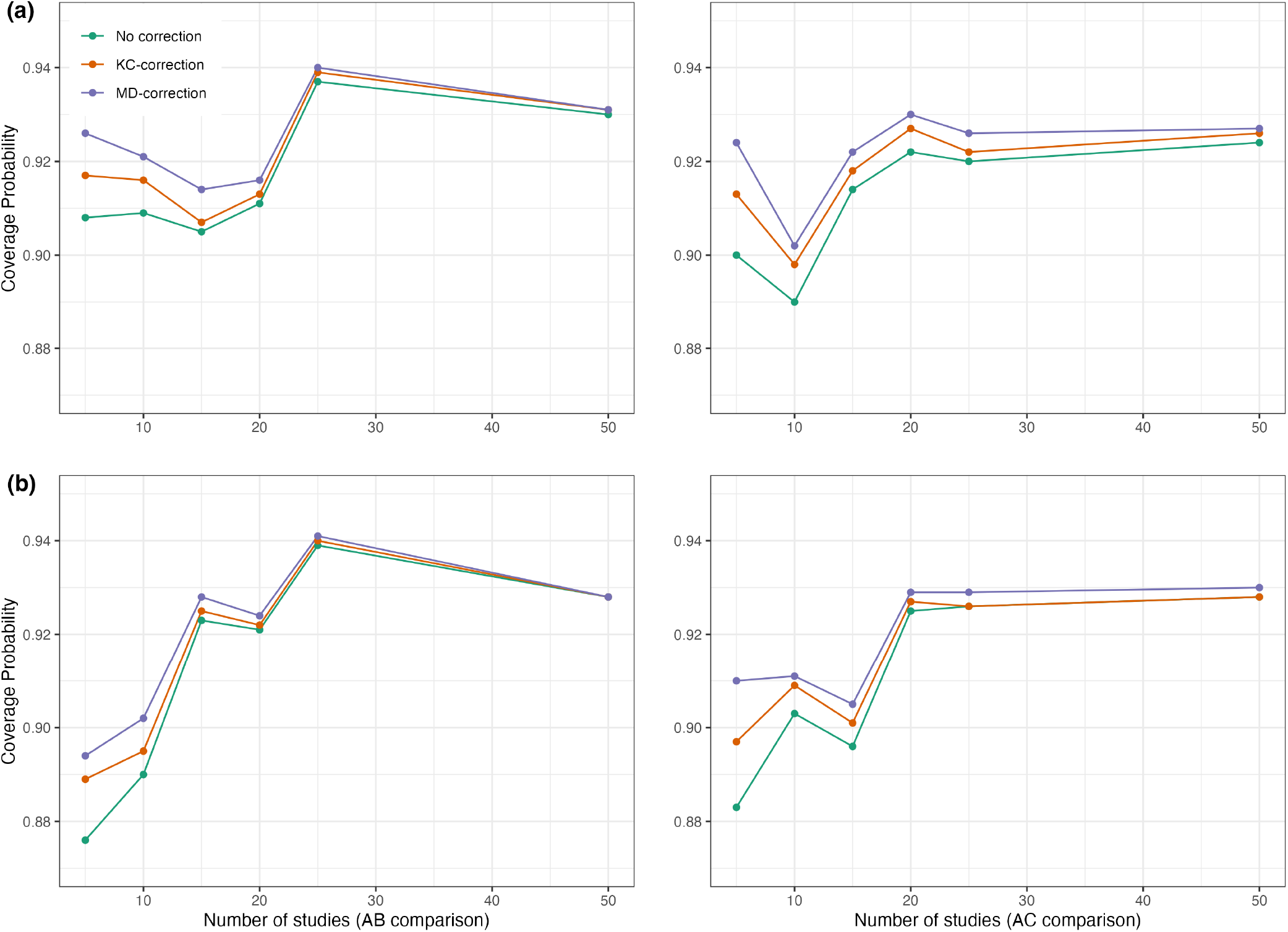
Coverage probabilities of estimated pooled treatment effects for comparisons between treatments *AB* and *AC* using the proposed method with and without the KC-corrected and MD-corrected sandwich variance estiamtors under (a) within-study correlation of 0.2; and (b) within-study correlation of 0.5.

On the other hand, results for the scenario with unequal between-study heterogeneity variance are provided in the lower panel of Table 1. The pooled estimates for *AB* and *AC* treatment comparisons were generally unbiased. As presented in the lower panel of Table 1, coverage probabilities were acceptable to good across most configurations, yielding coverage probabilities close to the nominal level of 95% (ranging from 89% to 94%). Similarly, variance estimates were adjusted using the KC-corrected and MD-corrected sandwich estimator for smaller studies, as illustrated in Tables S4 and S5 of the Supplementary Materials, respectively. As expected, coverage probabilities were slightly improved compared to results obtained from the proposed method without any corrections. In summary, simulation results suggested that the proposed method is robust to the magnitude of within-study correlations, regardless of whether the between-study heterogeneity variance is equal or unequal.

## 5 Data Application

In this section, we present the results of applying the proposed method to the two published NMAs, the primary open-angle glaucoma and the chronic prostatitis and chronic pelvic pain syndrome (CP/CPPS), as introduced in Section 2.

### 5.1 Application to primary open-angle glaucoma

The primary outcome of interest, in terms of IOP, was reported in a total of 22,656 patients across 125 studies, evaluating 4 classes of interventions, including *α*-2 adrenergic agonist, *β*-blocker, carbonic anhydrase inhibitor, and prostaglandin analogs (PAGs). For the IOP outcome, the within-study variances were generally much larger than the between-study variances; the between-study variance was estimated at approximately 0.49 using placebo as a reference. This was reflected by an *I*^2^ value of 43%, indicating that the total variation was not completely dominated by the between-study variation. Consequently, within-study correlations might lead to overestimated standard errors of pooled estimates for treatment comparisons if they were not properly accounted for in an NMA. The results of the standard NMA using the Lu and Ades’ approach (shown in the lower triangular matrix) and the proposed method (shown in the upper triangular matrix) without requiring knowledge of within-study correlations are presented in Figure S1 of the Supplementary Materials. The results of pairwise meta-analysis are provided in Figure S2 of the Supplementary Materials. As displayed in the upper triangular matrix of Figure S1, all active drugs were likely more effective in lowering IOP at 3 months compared to placebo, with mean differences in 3-month IOP ranging from *−*1.79 mmHg (95% CI, *−*2.65 to *−*0.94) to *−*5.53 mmHg (95% CI, *−*6.24 to *−*4.82). Moreover, bimatoprost showed the greatest reduction in 3-month IOP compared to placebo (mean difference = *−*5.53; 95% CI, *−*6.24 to *−*4.82), followed by travoprost (mean difference = *−*4.82; 95% CI, *−*5.51 to *−*4.12), latanoprost (mean difference = *−*4.61; 95% CI, *−*5.31 to *−*3.92), levobunolol (mean difference = *−*4.50; 95% CI, *−*5.68 to *−*3.32), taflurpost (mean difference = *−*3.93; 95% CI, *−*5.04 to *−*2.81), and so on. We noted that drugs within the PAGs class generally had similar effects on 3-month IOP reduction, except unoprostone (mean difference = *−*1.79; 95% CI, *−*2.65 to *−*0.94). Inconsistent results were found in several treatment comparisons when applying the standard NMA and the proposed method, including brinzolamide versus betaxolol (the standard NMA: mean difference = *−*0.85 with 95% CI, *−*1.71 to 0.00; the proposed: mean difference = *−*0.96 with 95% CI, *−*1.63 to *−*0.29), carteolol versus apraclonidine (the standard NMA: mean difference = *−*1.43 with 95% CI, *−*3.36 to 0.49; the proposed: mean difference = *−*1.47 with 95% CI, *−*2.69 to *−*0.24), tafluprost versus levobetaxolol (the standard NMA: mean difference = *−*1.47 with 95% CI, *−*2.81 to *−*0.12; the proposed: mean difference = *−*1.16 with 95% CI, *−*2.40 to 0.08), brinzolamide versus dorzolamide (the standard NMA: mean difference = *−*0.74 with 95% CI, *−*1.51 to 0.04; the proposed: mean difference = *−*0.73 with 95% CI, *−*1.30 to *−*0.16), and levobunolol versus carteolol (the standard NMA: mean difference = *−*1.09 with 95% CI, *−*2.15 to *−*0.03; the proposed: mean difference = *−*1.08 with 95% CI, *−*2.19 to 0.02).

Figure 4(a) illustrates the pooled estimates with corresponding 95% confidence intervals for all treatment comparisons using three approaches, including the pairwise meta-analysis, the standard NMA based on the Lu and Ades’ approach, as well as the proposed method. The pairwise meta-analysis showed the direct estimates from all available head-to-head comparisons. Obviously, it yielded wider 95% confidence intervals compared to the other two approaches due to that the indirect evidence was not borrowed into the analysis. The proposed method produced narrower 95% confidence intervals than the standard NMA approach for most of the treatment comparisons. Figure 5(a) displays a two-dimensional concordance plot of statistical significance represented by the Z value for both the standard NMA approach and the proposed method, where a Z value less than 1.96 equates to a p-value less than 0.05. A few points showed discordant evidence of treatment comparisons between the proposed method and the standard NMA. This discrepancy may be attributed to the fact that the proposed method took into account non-ignorable effects of within-study correlations, which affected the standard errors of the estimated pooled treatment effects. Figure S3(a) presents the treatment ranking based on the surface under the cumulative ranking curves (SUCRA)^48^ as mentioned in Section 3.3. A higher SUCRA score indicates a superior ranking for 3-month IOP reduction. Consequently, bimatoprost (SUCRA = 98.7%) had the highest SUCRA value for 3-month IOP reduction, followed by travoprost (SUCRA = 66.5%), latanoprost (SUCRA = 51.5%), and levobunolol (SUCRA = 42.5%).

**Figure 4:**
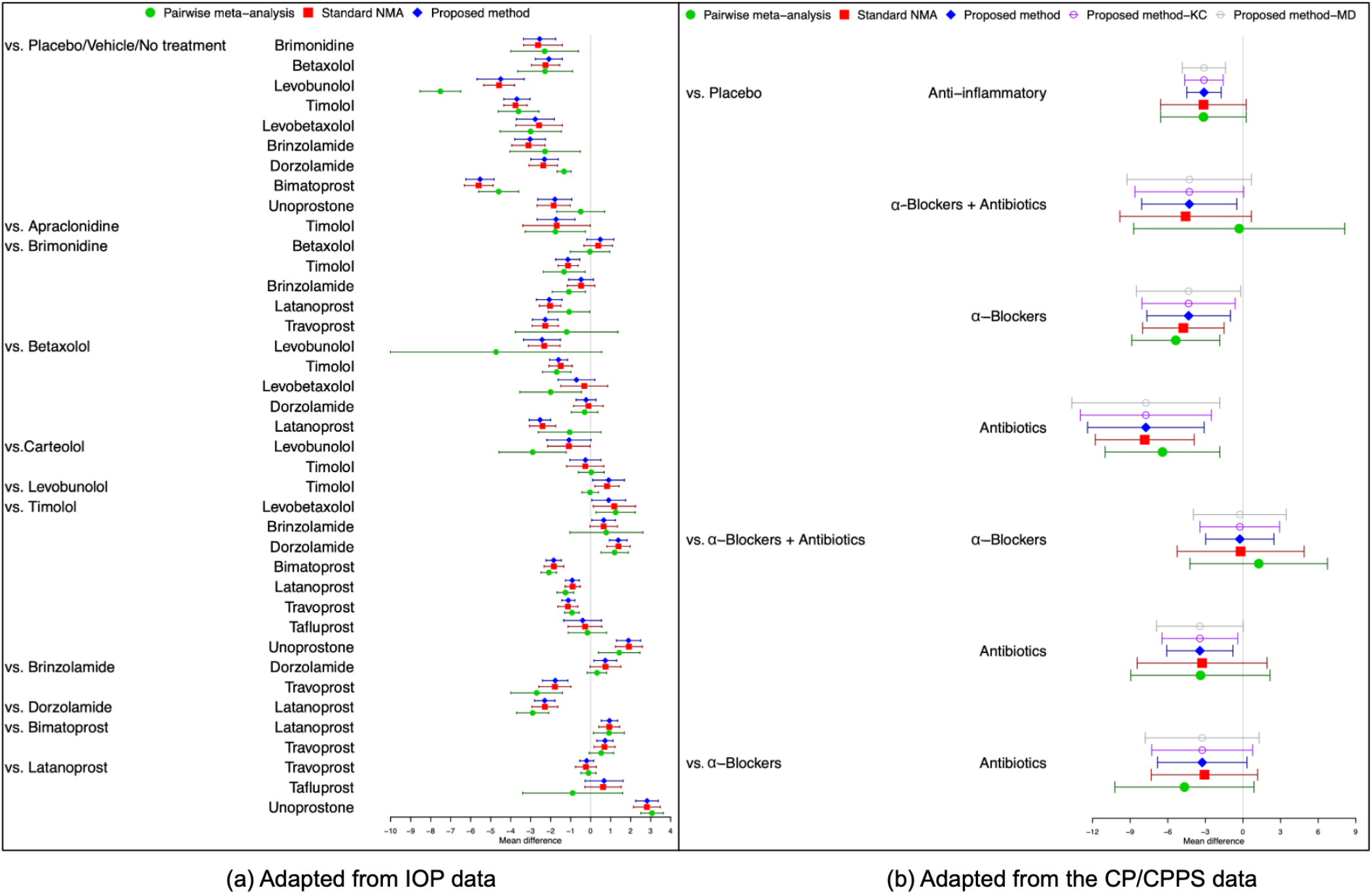
Comparisons of overall relative treatment estimates with 95% confidence intervals using the pairwise meta-analysis approach, the standard NMA based on the Lu and Ades’ approach, the proposed method without any corrections, and the proposed method with KC-corrected or MD-corrected sandwich variance estimators. Each node represents the pooled mean difference for the outcomes of interest.

**Figure 5:**
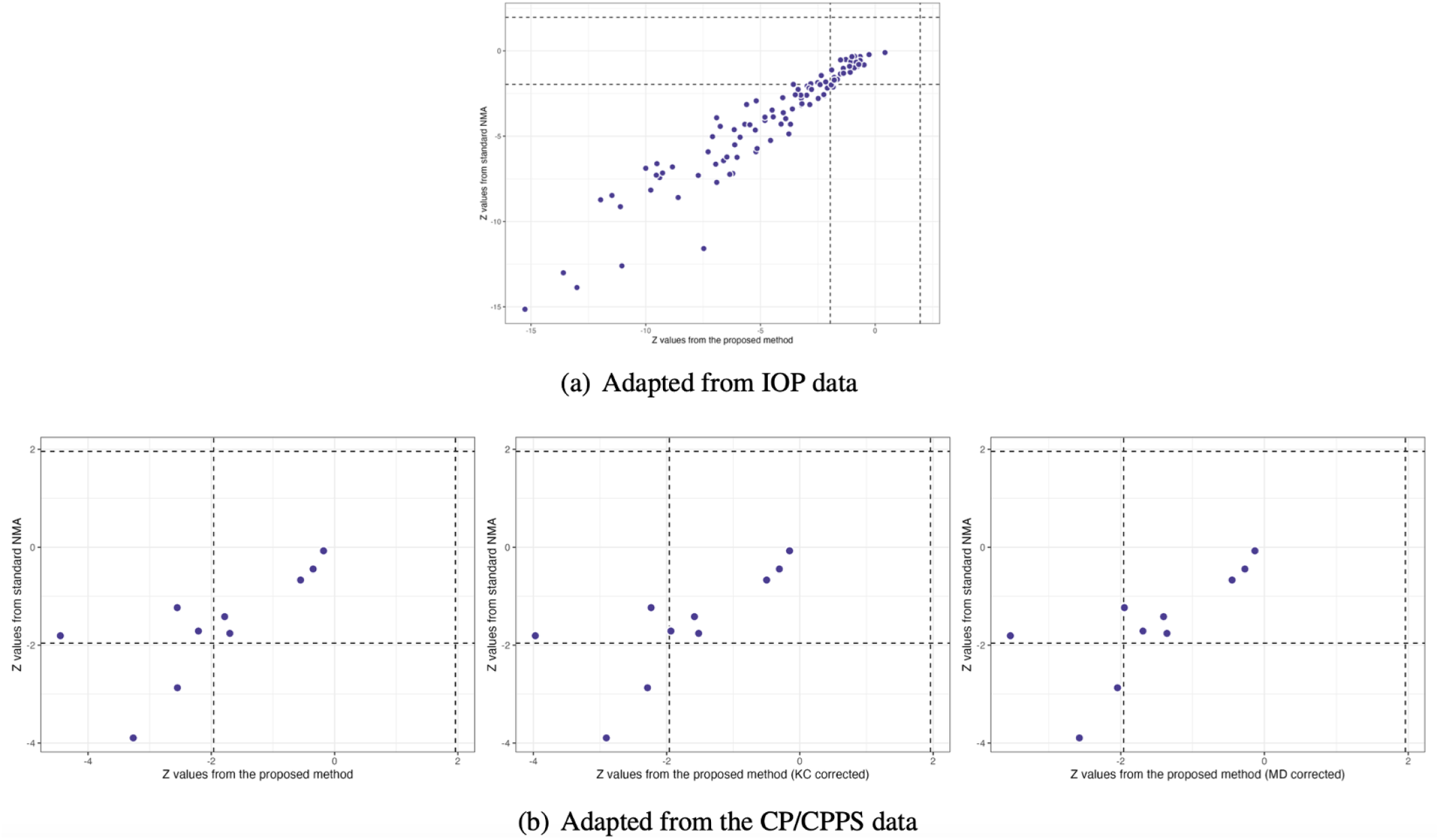
Comparisons of Z values using the standard NMA based on the Lu and Ades’ approach, the proposed method without any corrections, and the proposed method with KC-corrected or MD-corrected sandwich variance estimators, respectively

### 5.2 Application to chronic prostatitis and chronic pelvic pain syndrome

In this example, within-study variances are larger than between-study variances for the outcome of NIH-CPSI scores, and thus the effect of within-study correlations cannot be neglected. Due to the limited number of studies, we chose the KC-corrected and MD-corrected methods to adjust the sandwich-type variance estimations, following the rule of coefficient of variation ≤ 0.6. The NMA results using Lu and Ades’ approach and the proposed method without knowing within-study correlations are presented in Figure S4 of the Supplementary Materials. Upon re-analysis, in terms of NIH-CPSI scores, all active drugs appeared to be statistically significantly more effective than placebo, with mean differences of NIH-CPSI scores ranged from *−*7.75 (95% CI, *−*12.40 to *−*3.10) to *−*3.11 (95% CI, *−*4.48 to *−*1.74), as shown in Figure S4 of the Supplementary Materials. We found inconsistent results between the proposed method and the standard NMA method; *α*-blockers plus antibiotics (mean difference = *−*4.58, 95% CI: *−*9.82 to 0.67) and anti-inflammatory agents (mean difference = *−*3.15, 95% CI: *−*6.57 to 0.26) did not exhibit significantly greater efficacy than placebo with respect to NIH-CPSI scores when the standard NMA method was applied.

Moreover, the proposed method indicated that antibiotics alone was significantly more efficacious than *α*-blockers plus antibiotics (mean difference = *−*3.44; 95% CI, *−*6.08 to *−*0.80), a result not observed with the standard NMA method.

Figure 4(b) illustrates the overall relative estimates with 95% confidence intervals for all treatment comparisons using five different approaches, including the pairwise meta-analysis, the standard NMA method, and the proposed method with and without corrections. The results obtained from pairwise meta-analysis yielded wider 95% confidence intervals compared to all other NMA approaches. As expected, the proposed method had narrower 95% confidence intervals in contrast to the standard NMA using Lu and Ades’ approach. Furthermore, the proposed method without any corrections reported narrower 95% confidence intervals than the standard NMA approach. Corrections for the sandwich-type variance estimations using the KC-corrected and MD-corrected procedures resulted in slightly wider 95% confidence intervals than the proposed method without any corrections for some treatment comparisons. Figure 5(b) displays the concordance plot between the standard NMA approach and the proposed method with or without corrections. Several points showed discordant evidence of treatment comparisons between the proposed method and the standard NMA approach. This discrepancy may arise because the proposed method accounts for the unavailability of within-study correlations, which can affect the standard errors of estimated pooled treatment effects. Figure S3(b) displays treatment ranking with SUCRA. Overall, antibiotics (SUCRA = 94.6%) was ranked highest for the improvement of NIH-CPSI scores, followed by *α*-blockers plus antibiotics (SUCRA = 41.3%) and *α*-blockers (SUCRA = 42.4%).

## 6 Discussion

We proposed a composite likelihood-based approach to model univariate outcomes in NMAs. The proposed method helps obtain overall relative treatment effects in NMAs, even when within-study correlations are unavailable in the original articles. To the best of our knowledge, existing NMA approaches often make assumption about within-study correlations (known or zero), which can introduce bias if the true within-study correlations are non-negligible. Obtaining within-study correlations typically necessitates a joint analysis of individual participant-level data, often using bootstrapping methods.^68, 69^ However, this is seldom done unless specific questions about correlations between treatment comparisons are of interest in the included studies. Our proposed method has two key advantages. The first advantage is that the estimation and statistical inference of treatment effects are valid even if the correlation structure is misspecified. Simulation studies have shown that the proposed method provides nearly unbiased estimates and maintains reasonable coverage rates for 95% confidence intervals across scenarios with common or unequal between-study heterogeneity variances for treatment comparisons. Additionally, as illustrated in two applications, the proposed method is less prone to variance estimation issues than the standard NMA approach when total variation is not dominated by the between-study variation. The second advantage of the proposed method is its ability to reduce computational time by circumventing the need to estimate correlation parameters. This improvement is particularly evident when compared to currently available methods for NMAs, such as those implemented in the R packages ‘*gemtc*’ and ‘*netmeta*’ (see Figure 2 and Table S1 of the Supplementary Materials).

Nevertheless, there are two potential limitations to the proposed method. First, the proposed method focuses on contrast-based models in NMAs. While NMAs can also be performed using an arm-based approach, there are controversies in the literature regarding the differences between contrast-based and arm-based models.^20^ The arm-based approach offers a promising direction for future NMA modeling. It can potentially alleviate concerns about correlations among contrasts, especially in cases where treatment arm summaries are independent. Under these conditions, the arm-based approach yields the results consistent with the contrast-based method, requiring solely the standard errors of independent treatment summaries along with the inclusion of a fixed study main effect. Piepho and Madden^70^ demonstrated the practical application of an arm-based meta-analysis using SAS procedures GLIMMIX and BGLMM. Their work highlighted the effectiveness of this approach in circumventing the complexities associated with correlations among contrasts and maintaining concurrent control. However, a key criticism of the arm-based approach is that it might not fully preserve randomization within trials. This could potentially introduce bias in the estimated relative effects under certain scenarios, particularly if the assumption of transportable relative treatment effects is violated.^71^ Further research is needed to explore detailed formulations that can mitigate this issue and optimize the arm-based approach for NMAs.

Second, because the proposed method is constructed using a composite likelihood-based approach, the variance is estimated through a sandwich-type estimator. This estimator tends to underestimate the true variance, especially when the number of studies is small. This underestimation exacerbates the issue of under-coverage of confidence intervals and inflated type I error rates.^67^ To improve finite-sample variance estimation, we have applied the KC-corrected and MD-corrected sandwich variance estimators^63, 67^ in our simulation studies and the CP/CPPS application. As a result, the confidence intervals became slightly wider after applying these corrections, leading to improved coverage probabilities compared to the uncorrected method.

In conclusion, this work highlights the importance of considering non-ignorable within-study correlations in network meta-analyses. Ignoring these correlations, particularly when they are non-negligible, can lead to inaccurate standard errors for treatment effect estimates. The proposed composite likelihood-based approach offers an alternative for univariate NMAs when within-study correlation data is not available from original research articles. This method avoids the need for complex individual participant-level data analysis and maintains valid treatment effect estimation even with misspecified correlation structures. Additionally, it delivers significant computational efficiency gains compared to existing NMA methods.

## Data Availability

The network meta-analysis datasets described in Sections 2 and 5 were obtained from the following published studies: Li et al., Wang et al.,and Thakkinstian et al.

https://github.com/Penncil/xmeta/tree/master/R/CLNMA.equal.tau.R

## Data Availability Statement

The network meta-analysis datasets described in Sections 2 and 5 were obtained from the following published studies: Li et al.,^45^ Wang et al.,^46^ and Thakkinstian et al.^54^

