## Supplemental Tables and Figures for "Network meta-analysis made simple: a composite likelihood approach"

#### Appendix 1: Variance estimation

For the estimation of information matrix $\boldsymbol{I}$, we define:

$$\hat{\boldsymbol{I}} = -\frac{\partial^2 \ell(\boldsymbol{\eta})}{\partial \boldsymbol{\eta}^2}.$$

Meanwhile, define

$$\hat{\boldsymbol{\Lambda}} = \sum_{jj' \in \mathcal{T}} \sum_{i \in \mathcal{M}_{jj'}} S_{i,jj'} S_{i,jj'}^T,$$

where $S_{i,jj'}$ is denoted by

$$S_{i,jj'} = \frac{\partial \left[ \log \left( s_{i,jj'}^2 + \tau_{jj'}^2 \right) + \frac{(y_{i,jj'} - \mu_{jj'})^2}{s_{i,jj'}^2 + \tau_{jj'}^2} \right]}{\partial \eta}.$$

Then the variance of $\hat{\eta}$ can be estimated by the following sandwich-type estimator of form:

$$\hat{\boldsymbol{V}}(\hat{\boldsymbol{\eta}}) = \hat{\boldsymbol{I}}^{-1} \hat{\boldsymbol{\Lambda}} \hat{\boldsymbol{I}}^{-1}.$$

#### Appendix 2: Model with equal between-study variances

When we assume the same degree of heterogeneity for each contrast, i.e., $\tau_{jj'} = \tau$ for $\forall \tau_{jj'} \in \mathcal{T}$, the notations can be simplified as follows.

For the log-pseudolikelihood function $\ell(\boldsymbol{\eta})$, we simplify as

$$\ell(\boldsymbol{\eta}) = -\frac{1}{2} \sum_{jj' \in \mathcal{T}} \sum_{i \in \mathcal{M}_{jj'}} \left[ \log \left( s_{i,jj'}^2 + \tau^2 \right) + \frac{(y_{i,jj'} - \mu_{jj'})^2}{s_{i,jj'}^2 + \tau^2} \right].$$

For $\boldsymbol{H}^{(t)}$ and $\boldsymbol{\nu}^{(t)}$ in the system of linear equations $\boldsymbol{H}^{(t)} \boldsymbol{\mu} = \boldsymbol{\nu}^{(t)}$ by maximizing $\ell\left(\boldsymbol{\mu}, \tau^{2^{(t)}}\right)$ over $\boldsymbol{\mu}$, we have

$$\boldsymbol{H}^{(t)} = \begin{pmatrix} \sum_{j \in \mathcal{K} \setminus \{1\}} \sum_{i \in \mathcal{M}_{1j}} \omega_{i,1j}^{(t)} & -\sum_{i \in \mathcal{M}_{12}} \omega_{i,12}^{(t)} & \cdots & -\sum_{i \in \mathcal{M}_{1K}} \omega_{i,1K}^{(t)} \\ -\sum_{i \in \mathcal{M}_{12}} \omega_{i,12}^{(t)} & \sum_{j \in \mathcal{K} \setminus \{2\}} \sum_{i \in \mathcal{M}_{2j}} \omega_{i,2j}^{(t)} & \cdots & -\sum_{i \in \mathcal{M}_{2K}} \omega_{i,2K}^{(t)} \\ \vdots & \vdots & \ddots & \vdots \\ -\sum_{i \in \mathcal{M}_{1K}} \omega_{i,1K}^{(t)} & -\sum_{i \in \mathcal{M}_{2K}} \omega_{i,2K}^{(t)} & \cdots & \sum_{j \in \mathcal{K} \setminus \{K\}} \sum_{i \in \mathcal{M}_{Kj}} \omega_{i,Kj}^{(t)} \end{pmatrix};$$

$$\boldsymbol{v}^{(t)} = \begin{pmatrix} \sum_{j\in\mathcal{K}\setminus\{1\}} \sum_{i\in\mathcal{M}_{1j}} y_{i,j1}\omega_{i,1j}^{(t)} \\ \sum_{j\in\mathcal{K}\setminus\{2\}} \sum_{i\in\mathcal{M}_{2j}} y_{i,j2}\omega_{i,2j}^{(t)} \\ \vdots \\ \sum_{j\in\mathcal{K}\setminus\{K\}} \sum_{i\in\mathcal{M}_{K}} y_{i,jK}\omega_{i,Kj}^{(t)} \end{pmatrix}; \text{ and } \omega_{i,jj'}^{(t)} = \left(s_{i,jj'}{}^2 + \tau^{2(t)}\right)^{-1}.$$

To estimate the variance of the estimator from the simplified situation, we have

$$\tilde{\boldsymbol{I}} = -\frac{\partial^2 \ell(\boldsymbol{\eta})}{\partial \boldsymbol{\eta}^2}$$

and

$$\tilde{\boldsymbol{\Lambda}} = \sum_{jj'\in\mathcal{T}} \sum_{i\in\mathcal{M}_{jj'}} S_{i,jj'} S_{i,jj'}^T,$$

where $S_{i,jj'}$ is denoted by

$$S_{i,jj'} = \frac{\partial \left[\log\left(s_{i,jj'}^2 + \tau^2\right) + \frac{(y_{i,jj'} - \mu_{jj'})^2}{s_{i,jj'}^2 + \tau^2}\right]}{\partial \eta}.$$

Then the variance of $\tilde{\eta}$ can be estimated by the following sandwich-type estimator of form:

$$\tilde{\boldsymbol{V}}(\tilde{\boldsymbol{\eta}}) = \tilde{\boldsymbol{I}}^{-1} \tilde{\boldsymbol{\Lambda}} \tilde{\boldsymbol{I}}^{-1}.$$

### Appendix 3: Proof of convergence for the Algorithm 1

**Regularity Conditions**

(C1). The parameter space $\boldsymbol{\Theta}$ contains an open set of which the true parameter value $\boldsymbol{\eta}^*$ is an interior point.

(C2). The proportion of studies that report the estimated treatment effects and standard errors of every treatment comparison converges to a constant as the number of included studies $m$ goes to infinity.

(C3). The marginal population risk, defined as

$$l^{jj'}(\boldsymbol{\eta}) = -\mathbb{E} \sum_{i\in\mathcal{M}_{jj'}} \left[\log\left(s_{i,jj'}^2 + \tau^2\right) + \frac{(y_{i,jj'} - \mu_{jj'})^2}{s_{i,jj'}^2 + \tau^2}\right],$$

has an unique maximizer, which is the true parameter value $\boldsymbol{\eta}^*$.

(C4). The first, second and third order derivatives of $\ell(\boldsymbol{\eta})$ are all bounded.

(C5). The limit of the information matrix is positive definite, and for any $\boldsymbol{\eta} \in \boldsymbol{\Theta}$, the minimal eigenvalue $\lambda_{\min}$ of the limiting information matrix satisfies $\lambda_{\min} > c > 0$ for some constant $c > 0$.

**Lemma S1.** Denote $\ell_m(\boldsymbol{\eta}) = \ell(\boldsymbol{\eta})/m$ as the normalized log-pseudolikelihood function. For any given number of iteration $t$ and sufficiently large $m$, there exist some constant $C > 0$ such that the obtained $\boldsymbol{\eta}^{(t)}$ from Algorithm 1 satisfies

$$|\ell_m(\boldsymbol{\eta}^{(t)}) - \ell_m^*| \leq \frac{C}{t},$$

where $\ell_m^* = \max_{\boldsymbol{\eta}} \ell_m(\boldsymbol{\eta})$.

*Proof.* Denote $I(\boldsymbol{\eta}) = \lim_{m\to\infty} -\mathbb{E}\frac{\partial^2 \ell_m(\boldsymbol{\eta})}{\partial \boldsymbol{\eta}^2}$ to be the limit of the information matrix $\boldsymbol{I}(\boldsymbol{\eta})$. From assumption (C4) and (C5), for sufficiently large $m$, we have

$$\lambda_{\min} \cdot \left(-\frac{\partial^2 \ell_m(\boldsymbol{\eta})}{\partial \boldsymbol{\eta}^2}\right) \geq \lambda_{\min} \cdot I(\boldsymbol{\eta}) - \lambda_{\max} \cdot \left(-\frac{\partial^2 \ell_m(\boldsymbol{\eta})}{\partial \boldsymbol{\eta}^2} - I(\boldsymbol{\eta})\right) \geq c - \frac{c}{2} = \frac{c}{2},$$

which implies that $\ell_m(\boldsymbol{\eta})$ is a convex function. Apply Theorem 3.2 in **?** and we finish the proof.

**Lemma S2.** For any given number of iteration $t$ and sufficiently large $m$, there exist some constant $C > 0$ such that the obtained $\boldsymbol{\eta}^{(t)}$ from Algorithm 1 satisfies

$$(\ell_m(\boldsymbol{\eta}^{(t+1)}) - \ell_m^*)^2 \leq C||\tau^{2^{(t+1)}} - \tau^{2^{(t)}}||_2.$$

*Proof.* We have

$$(\ell_m(\boldsymbol{\mu}^{(t+1)}, \tau^{2^{(t+1)}}) - \ell_m^*)^2 \leq \tilde{C}(\ell_m(\boldsymbol{\mu}^{(t)}, \tau^{2^{(t+1)}}) - \ell_m(\boldsymbol{\mu}^{(t)}, \tau^{2^{(t)}})) \leq C||\tau^{2^{(t+1)}} - \tau^{2^{(t)}}||_2,$$

where the first inequality can be obtained by applying Lemma 3.4 in **?**, and the boundedness of gradients leads to the second inequality. In this case, we can say that we are close to the global minimum when $||\tau^{2^{(t+1)}} - \tau^{2^{(t)}}||_2$ is small.

### Appendix 4: Testing inconsistency

NMAs rely on two key assumptions, including transitivity and consistency. Indeed, the estimates from NMAs are valid only under the assumption of consistency **??**. Unlike the assumption of transitivity, inconsistency can be always evaluated statistically, which is known as inconsistency when the direct and indirect evidence are not in agreement. Along with the work in **?**, the inconsistency model is depicted by the following equation

$$\mu_{BC} = \mu_{AC} - \mu_{AB} + \phi.$$

Here, $\phi$ represents an inconsistency parameter between the indirect evidence from $BC$ treatment comparison and the direct evidence from $AB$ and $AC$ treatment comparisons. One can evaluate and test whether the consistency assumption is satisfied with $\phi = 0$.

### Appendix 5: A working example

This working example compares the restricted maximum likelihood (REML) and the generalized estimating equations (GEE) under a consistency constraint on the pairwise means. For illustration purposes, we considered a 3-arm example. Specifically, suppose that we have an NMA consisting of three treatments: $A$, $B$, and $C$, with treatment $A$ assumed to be the reference and included in every trial. The set of observed treatment comparisons is $\mathcal{T} = \{AB, BC, AC\}$. Let $\mathcal{M}_{jj'}$ be the subset of studies that report effect sizes and standard errors of the outcome for the treatment comparison between $j$ and $j'$. Recall that the log-likelihood function of the multivariate random-effects model $\ell(\boldsymbol{\eta}, \boldsymbol{\tau}^2)$ is written as

$$\ell(\boldsymbol{\mu}, \boldsymbol{\tau}^2) = -\frac{1}{2} \sum_{i \in \mathcal{M}_{jj'}} \left\{\log|\Sigma + \Omega_i| + (\boldsymbol{y}_i - \boldsymbol{\mu})^T (\Sigma + \Omega_i)^{-1} (\boldsymbol{y}_i - \boldsymbol{\mu}) + \log(2\pi)\right\}.$$

The REML approach involves adding an extra term to the log-likelihood function $\ell(\boldsymbol{\mu}, \boldsymbol{\tau}^2)$ with $-\frac{1}{2}\sum_{i\in\mathcal{M}_{jj'}} \log|(\Sigma+\Omega_i)^{-1}|$, that is,

$$\ell_{REML}(\boldsymbol{\mu}, \boldsymbol{\tau}^2) = -\frac{1}{2}\sum_{i\in\mathcal{M}_{jj'}} \left\{\log|\Sigma+\Omega_i| + (\boldsymbol{y}_i-\boldsymbol{\mu})^T(\Sigma+\Omega_i)^{-1}(\boldsymbol{y}_i-\boldsymbol{\mu}) + \log(2\pi) + \log|(\Sigma+\Omega_i)^{-1}|\right\}.$$

The score function of the REML log-likelihood for $\boldsymbol{\mu}$ is given by

$$\frac{\partial \ell_{REML}(\boldsymbol{\mu}, \boldsymbol{\tau}^2)}{\partial \boldsymbol{\mu}} = \sum_{i\in\mathcal{M}_{jj'}} (\boldsymbol{\Sigma}+\boldsymbol{\Omega}_i)^{-1}(y_i-\boldsymbol{\mu}) = 0,$$

and the second derivative is written as

$$\frac{\partial^2 \ell_{REML}(\boldsymbol{\mu}, \boldsymbol{\tau}^2)}{\partial \boldsymbol{\mu}^2} = -\sum_{i\in\mathcal{M}_{jj'}} (\boldsymbol{\Sigma}+\boldsymbol{\Omega}_i)^{-1}.$$

We note that the REML approach yields the same asymptotic distribution as the maximum likelihood (ML) results. The system of linear equations derived from the score equations can be iteratively solved. That is, by maximizing $\ell_{REML}\left(\boldsymbol{\mu}, \boldsymbol{\tau}_{jj'}^{2(t)}\right)$ over $\boldsymbol{\mu}$, one can obtain the estimates of $\boldsymbol{\mu}$ from $\sum_{i\in\mathcal{M}_{jj'}}(\boldsymbol{\Sigma}+\boldsymbol{\Omega}_i)^{-1(t)}\boldsymbol{y}_i = \sum_{i\in\mathcal{M}_{jj'}}(\boldsymbol{\Sigma}+\boldsymbol{\Omega}_i)^{-1(t)}\boldsymbol{\mu}$, where

$$(\boldsymbol{\Sigma}+\boldsymbol{\Omega}_i)^{-1(t)} = \begin{pmatrix} \sum_{i\in\mathcal{M}_{AB}}\omega_{i,AB}^{(t)} + \sum_{i\in\mathcal{M}_{BC}}\omega_{i,BC}^{(t)} & -\sum_{i\in\mathcal{M}_{BC}}\omega_{i,BC}^{(t)} \\ -\sum_{i\in\mathcal{M}_{BC}}\omega_{i,BC}^{(t)} & \sum_{i\in\mathcal{M}_{AC}}\omega_{i,AC}^{(t)} + \sum_{i\in\mathcal{M}_{BC}}\omega_{i,BC}^{(t)} \end{pmatrix};$$

$$(\boldsymbol{\Sigma}+\boldsymbol{\Omega}_i)^{-1(t)}\boldsymbol{y}_i = \begin{pmatrix} \sum_{i\in\mathcal{M}_{AB}} y_{i,AB}\omega_{i,AB}^{(t)} - \sum_{i\in\mathcal{M}_{BC}} y_{i,BC}\omega_{i,BC}^{(t)} \\ \sum_{i\in\mathcal{M}_{AC}} y_{i,AC}\omega_{i,AC}^{(t)} + \sum_{i\in\mathcal{M}_{BC}} y_{i,BC}\omega_{i,BC}^{(t)} \end{pmatrix}; \text{ and } \omega_{i,jj'}^{(t)} = \left(s_{i,jj'}^2 + \tau_{jj'}^{2(t)}\right)^{-1}.$$

Regarding GEE, it was obtained by solving the following estimating equations for $\boldsymbol{\mu}$:

$$\sum_{i\in\mathcal{M}_{jj'}} \left\{\frac{\partial E(\boldsymbol{y}_i)}{\partial \boldsymbol{\mu}}\right\}^T \boldsymbol{V}_i^{-1}(\boldsymbol{y}_i-\boldsymbol{\mu}) = 0.$$

By iteratively least squares method, one can update $\boldsymbol{\mu}$ through the following equation until convergence

$$\boldsymbol{\mu}^{(t+1)} = \boldsymbol{\mu}^{(t)} + \left(\sum_{i\in\mathcal{M}_{jj'}} \left\{\frac{\partial E(\boldsymbol{y}_i)}{\partial \boldsymbol{\mu}}\right\}^{T(t)} \boldsymbol{V}_i^{(t)^{-1}} \left\{\frac{\partial E(\boldsymbol{y}_i)}{\partial \boldsymbol{\mu}}\right\}^{(t)}\right)^{-1} \sum_{i\in\mathcal{M}_{jj'}} \left\{\frac{\partial E(\boldsymbol{y}_i)}{\partial \boldsymbol{\mu}}\right\}^{T(t)} \boldsymbol{V}_i^{(t)^{-1}}(\boldsymbol{y}_i - \boldsymbol{\mu}^{(t)}),$$

where $\boldsymbol{y}_i = (y_{i,AB}, y_{i,BC}, y_{i,AC})^T$, $E(\boldsymbol{y}_i) = \boldsymbol{\mu} = (\mu_{i,AB}, \mu_{i,BC}, \mu_{i,AC})^T$.

### Appendix 6: Additional results for simulation studies and applications

Table S1 summarized the computational time for the two existing methods, implemented in 'gemtc' and 'netmeta', along with the proposed method. Tables S2 and S3 summarized the performance of the proposed method with KC-based correction and MD-based correction separately, under the scenario of common between-study heterogeneity variance ($\tau_{AB} = \tau_{AC} = 0.5$, $\mu_{AB} = -2$, and $\mu_{AC} = -4$). Tables S4 and S5 summarized the simulation results based on the proposed method with KC-based and MD-based corrections, respectively, in a scenario with unequal between-study heterogeneity variance ($\tau_{AB} = 0.7$, $\tau_{AC} = 0.5$, $\mu_{AB} = -2$, and $\mu_{AC} = -4$). Figure S2 showed the results fitted by the proposed method and the pairwise meta-analysis for the IOP data. Figure S3 illustrated the results of treatment ranking based on SUCRA for both the IOP data (Figure S3(a)) and the chronic prostatitis and chronic pelvic pain syndrome data (Figure S3(b)). Figure S4 reported the results obtained from the standard NMA using Lu and Ades' approach, along with the results from the proposed method, for the chronic prostatitis and chronic pelvic pain syndrome data.

Web Table S1: Comparison of computational time in seconds for the existing R packages and the proposed method using a simulated dataset.

| Number of treatments under comparisons | Total number of studies | Computational time (seconds) | | |
|---|---|---|---|---|
| | | **gemtc** [Bayesian approach] | **netmeta** [Frequentist approach] | **Proposed** |
| 5 | 10 | 1.581 | 1.814 | 0.411 |
| | 20 | 2.399 | 2.157 | 0.385 |
| | 30 | 4.828 | 2.476 | 0.453 |
| | 40 | 5.617 | 2.884 | 0.561 |
| | 50 | 8.612 | 2.916 | 0.523 |
| 10 | 40 | 9.318 | 7.392 | 3.395 |
| | 60 | 17.481 | 11.721 | 4.166 |
| | 80 | 28.476 | 17.463 | 4.201 |
| | 100 | 49.044 | 40.682 | 5.177 |
| | 120 | 61.755 | 69.143 | 5.670 |
| 15 | 120 | 12.967 | 9.156 | 34.013 |
| | 150 | 38.263 | 35.275 | 26.756 |
| | 180 | 70.848 | 139.04 | 20.091 |
| | 210 | 95.597 | 321.474 | 24.637 |
| | 240 | 138.93 | 858.16 | 37.626 |

Web Table S2: Summary of 1,000 simulations with m = 5, 10, 15, 20, 25 and 50: bias (Bias), empirical standard error (ESE), model-based standard error (MBSE) and coverage probability (CP) of pooled estimates of $AB$ and $AC$ treatment comparisons. The data-generation mechanism was through a common between-study heterogeneity variance with $\tau_{AB} = \tau_{AC} = 0.5$, $\mu_{AB} = -2$, and $\mu_{AC} = -4$. Results were based on the proposed method with KC-correction.

| Within-study correlation | Number of studies | AB comparison after KC-correction | | | | AC comparison after KC-correction | | | |
|---|---|---|---|---|---|---|---|---|---|
| | | Bias | ESE | MBSE | CP | Bias | ESE | MBSE | CP |
| 0.2 | 5 | 0.0118 | 0.3102 | 0.2962 | 0.917 | -0.0038 | 0.3261 | 0.2974 | 0.913 |
| | 10 | 0.0044 | 0.2361 | 0.2110 | 0.916 | 0.0037 | 0.2468 | 0.2139 | 0.898 |
| | 15 | -0.0002 | 0.1925 | 0.1732 | 0.907 | -0.0010 | 0.1932 | 0.1733 | 0.918 |
| | 20 | -0.0018 | 0.1646 | 0.1496 | 0.913 | -0.0012 | 0.1647 | 0.1500 | 0.927 |
| | 25 | 0.0008 | 0.1428 | 0.1342 | 0.939 | -0.0003 | 0.1456 | 0.1341 | 0.922 |
| | 50 | 0.0012 | 0.1030 | 0.0948 | 0.931 | 0.0025 | 0.1059 | 0.0949 | 0.926 |
| 0.5 | 5 | -0.0079 | 0.3456 | 0.2875 | 0.889 | -0.0026 | 0.3284 | 0.2892 | 0.897 |
| | 10 | 0.0218 | 0.2347 | 0.2044 | 0.895 | 0.013 | 0.2279 | 0.2041 | 0.909 |
| | 15 | 0.0013 | 0.1817 | 0.1687 | 0.925 | -0.0015 | 0.1948 | 0.1676 | 0.901 |
| | 20 | -0.0011 | 0.1594 | 0.1457 | 0.922 | -0.0087 | 0.1572 | 0.1456 | 0.927 |
| | 25 | -0.0033 | 0.1386 | 0.1300 | 0.940 | -0.0073 | 0.1410 | 0.1302 | 0.926 |
| | 50 | 0.0064 | 0.1007 | 0.0920 | 0.928 | 0.0031 | 0.0998 | 0.0920 | 0.928 |

Web Table S3: Summary of 1,000 simulations with m = 5, 10, 15, 20, 25 and 50: bias (Bias), empirical standard error (ESE), model-based standard error (MBSE) and coverage probability (CP) of pooled estimates of $AB$ and $AC$ treatment comparisons. The data-generation mechanism was through a common between-study heterogeneity variance with $\tau_{AB} = \tau_{AC} = 0.5$, $\mu_{AB} = -2$, and $\mu_{AC} = -4$. Results were based on the proposed method with MD-correction.

| Within-study correlation | Number of studies | AB comparison after MD-correction | | | | AC comparison after MD-correction | | | |
|---|---|---|---|---|---|---|---|---|---|
| | | Bias | ESE | MBSE | CP | Bias | ESE | MBSE | CP |
| 0.2 | 5 | 0.0118 | 0.3102 | 0.3067 | 0.926 | -0.0038 | 0.3261 | 0.3080 | 0.924 |
| | 10 | 0.0044 | 0.2361 | 0.2147 | 0.921 | 0.0037 | 0.2468 | 0.2176 | 0.902 |
| | 15 | -0.0002 | 0.1925 | 0.1752 | 0.914 | -0.0010 | 0.1932 | 0.1753 | 0.922 |
| | 20 | -0.0018 | 0.1646 | 0.1509 | 0.916 | -0.0012 | 0.1647 | 0.1512 | 0.930 |
| | 25 | 0.0008 | 0.1428 | 0.1351 | 0.940 | -0.0003 | 0.1456 | 0.1350 | 0.926 |
| | 50 | 0.0012 | 0.1030 | 0.0951 | 0.931 | 0.0025 | 0.1059 | 0.0952 | 0.927 |
| 0.5 | 5 | -0.0079 | 0.3456 | 0.2977 | 0.894 | -0.0026 | 0.3284 | 0.2994 | 0.910 |
| | 10 | 0.0218 | 0.2347 | 0.2079 | 0.902 | 0.013 | 0.2279 | 0.2076 | 0.911 |
| | 15 | 0.0013 | 0.1817 | 0.1706 | 0.928 | -0.0015 | 0.1948 | 0.1695 | 0.905 |
| | 20 | -0.0011 | 0.1594 | 0.1469 | 0.924 | -0.0087 | 0.1572 | 0.1468 | 0.929 |
| | 25 | -0.0033 | 0.1386 | 0.1309 | 0.941 | -0.0073 | 0.1410 | 0.1310 | 0.929 |
| | 50 | 0.0064 | 0.1007 | 0.0923 | 0.928 | 0.0031 | 0.0998 | 0.0923 | 0.930 |

Web Table S4: Summary of 1,000 simulations with m = 5, 10, 15, 20, 25 and 50: bias (Bias), empirical standard error (ESE), model-based standard error (MBSE) and coverage probability (CP) of pooled estimates of $AB$ and $AC$ treatment comparisons. The data-generation mechanism was through unequal between-study heterogeneity variances with $\tau_{AB} = 0.7$, $\tau_{AC} = 0.5$, $\mu_{AB} = -2$, and $\mu_{AC} = -4$. Results were based on the proposed method with KC-correction.

| Within-study correlation | Number of studies | AB comparison after KC-correction | | | | AC comparison after KC-correction | | | |
|---|---|---|---|---|---|---|---|---|---|
| | | Bias | ESE | MBSE | CP | Bias | ESE | MBSE | CP |
| 0.2 | 5 | -0.0100 | 0.3473 | 0.3192 | 0.905 | 0.0077 | 0.3393 | 0.3072 | 0.910 |
| | 10 | 0.0108 | 0.2487 | 0.2262 | 0.919 | -0.0074 | 0.2396 | 0.2175 | 0.915 |
| | 15 | -0.0003 | 0.2032 | 0.1864 | 0.927 | 0.0012 | 0.1888 | 0.1782 | 0.931 |
| | 20 | -0.0059 | 0.1744 | 0.1603 | 0.931 | -0.0064 | 0.1691 | 0.1535 | 0.922 |
| | 25 | -0.0019 | 0.1565 | 0.1438 | 0.919 | 0.0021 | 0.1484 | 0.1373 | 0.919 |
| | 50 | -0.0017 | 0.1146 | 0.1021 | 0.919 | -0.0016 | 0.0996 | 0.0976 | 0.943 |
| 0.5 | 5 | 0.0057 | 0.355 | 0.3129 | 0.908 | 0.0066 | 0.321 | 0.2985 | 0.921 |
| | 10 | -0.0024 | 0.2454 | 0.2212 | 0.915 | -0.0013 | 0.2265 | 0.2124 | 0.932 |
| | 15 | 0.0081 | 0.2086 | 0.1806 | 0.903 | 0.0134 | 0.1932 | 0.1731 | 0.914 |
| | 20 | 0.0061 | 0.1786 | 0.1558 | 0.904 | 0.0050 | 0.1660 | 0.1492 | 0.916 |
| | 25 | 0.0001 | 0.1610 | 0.1403 | 0.905 | 0.0011 | 0.1447 | 0.1341 | 0.913 |
| | 50 | 0.0057 | 0.1092 | 0.0990 | 0.916 | 0.0076 | 0.1012 | 0.0949 | 0.933 |

Web Table S5: Summary of 1,000 simulations with m = 5, 10, 15, 20, 25 and 50: bias (Bias), empirical standard error (ESE), model-based standard error (MBSE) and coverage probability (CP) of pooled estimates of $AB$ and $AC$ treatment comparisons. The data-generation mechanism was through unequal between-study heterogeneity variances with $\tau_{AB} = 0.7$, $\tau_{AC} = 0.5$, $\mu_{AB} = -2$, and $\mu_{AC} = -4$. Results were based on the proposed method with MD-correction.

| Within-study | Number | AB comparison after MD-correction | | | | AC comparison after MD-correction | | | |
|---|---|---|---|---|---|---|---|---|---|
| correlation | of studies | Bias | ESE | MBSE | CP | Bias | ESE | MBSE | CP |
| 0.2 | 5 | -0.0100 | 0.3473 | 0.3305 | 0.912 | 0.0077 | 0.3393 | 0.3181 | 0.919 |
| | 10 | 0.0108 | 0.2487 | 0.2301 | 0.923 | -0.0074 | 0.2396 | 0.2212 | 0.918 |
| | 15 | -0.0003 | 0.2032 | 0.1886 | 0.929 | 0.0012 | 0.1888 | 0.1802 | 0.934 |
| | 20 | -0.0059 | 0.1744 | 0.1617 | 0.933 | -0.0064 | 0.1691 | 0.1548 | 0.924 |
| | 25 | -0.0019 | 0.1565 | 0.1448 | 0.921 | 0.0021 | 0.1484 | 0.1382 | 0.922 |
| | 50 | -0.0017 | 0.1146 | 0.1024 | 0.919 | -0.0016 | 0.0996 | 0.0980 | 0.944 |
| 0.5 | 5 | 0.0057 | 0.355 | 0.3239 | 0.919 | 0.0066 | 0.321 | 0.3090 | 0.929 |
| | 10 | -0.0024 | 0.2454 | 0.2250 | 0.919 | -0.0013 | 0.2265 | 0.2161 | 0.932 |
| | 15 | 0.0081 | 0.2086 | 0.1827 | 0.912 | 0.0134 | 0.1932 | 0.1750 | 0.919 |
| | 20 | 0.0061 | 0.1786 | 0.1571 | 0.908 | 0.0050 | 0.1660 | 0.1505 | 0.921 |
| | 25 | 0.0001 | 0.1610 | 0.1413 | 0.907 | 0.0011 | 0.1447 | 0.1350 | 0.916 |
| | 50 | 0.0057 | 0.1092 | 0.0993 | 0.916 | 0.0076 | 0.1012 | 0.0953 | 0.933 |

| Bimatoprost | Travoprost | Latanoprost | Levobunolol | Tafluprost | Timolol | Carteolol | Brinzolamide | Brimonidine | Levobetaxolol | Dorzolamide | Betaxolol | Apraclonidine | Unoprostone | Placebo |
|---|---|---|---|---|---|---|---|---|---|---|---|---|---|---|
| Bimatoprost | -0.72 (-1.12, -0.31) | -0.94 (-1.35, -0.53) | -1.03 (-1.93, -0.13) | -1.59 (-2.56, -0.61) | -1.85 (-2.23, -1.47) | -2.11 (-2.97, -1.25) | -2.49 (-3.16, -1.82) | -2.98 (-3.66, -2.30) | -2.75 (-3.66, -1.83) | -3.22 (-3.77, -2.67) | -3.45 (-4.02, -2.89) | -3.58 (-4.59, -2.56) | -3.74 (-4.40, -3.08) | -5.53 (-6.24, -4.82) |
| -0.69 (-1.22, -0.17) | Travoprost | -0.20 (-0.55, 0.15) | -0.32 (-1.19, 0.56) | -0.87 (-1.83, 0.08) | -1.12 (-1.44, -0.79) | -1.39 (-2.23, -0.55) | -1.77 (-2.41, -1.14) | -2.27 (-2.91, -1.63) | -2.03 (-2.92, -1.13) | -2.50 (-3.02, -1.99) | -2.73 (-3.27, -2.20) | -2.86 (-3.86, -1.86) | -3.02 (-3.65, -2.39) | -4.82 (-5.51, -4.12) |
| -0.93 (-1.45, -0.41) | -0.24 (-0.76, 0.28) | Latanoprost | -0.12 (-0.99, 0.76) | -0.67 (-1.62, 0.28) | -0.92 (-1.26, -0.57) | -1.19 (-2.03, -0.35) | -1.57 (-2.21, -0.93) | -2.07 (-2.71, -1.43) | -1.83 (-2.72, -0.93) | -2.30 (-2.81, -1.79) | -2.53 (-3.07, -2.00) | -2.66 (-3.66, -1.65) | -2.82 (-3.38, -2.25) | -4.61 (-5.31, -3.92) |
| -1.02 (-1.79, -0.24) | -0.32 (-1.10, 0.45) | -0.08 (-0.79,0.62) | Levobunolol | -0.56 (-1.77, 0.65) | -0.90 (-1.69, -0.10) | -1.08 (-2.19, 0.02) | -1.47 (-2.48, -0.46) | -1.95 (-2.96, -0.93) | -1.72 (-2.90, -0.54) | -2.20 (-3.15, -1.26) | -2.43 (-3.36, -1.51) | -2.55 (-3.79, -1.31) | -2.72 (-3.74, -1.69) | -4.50 (-5.68, -3.32) |
| -1.55 (-2.52, -0.58) | -0.86 (-1.83, 0.12) | -0.62 (-1.53,0.29) | -0.53 (-1.57, 0.50) | Tafluprost | -0.40 (-1.35, 0.54) | -0.52 (-1.72, 0.67) | -0.91 (-1.99, 0.17) | -1.37 (-2.45, -0.29) | -1.16 (-2.40, 0.08) | -1.65 (-2.65, -0.64) | -1.87 (-2.89, -0.86) | -1.99 (-3.31, -0.68) | -2.16 (-3.24, -1.07) | -3.93 (-5.04, -2.81) |
| -1.84 (-2.33, -1.34) | -1.14 (-1.64, -0.64) | -0.90 (-1.28, -0.52) | -0.82 (-1.42, -0.21) | -0.28 (-1.13,0.56) | Timolol | -0.26 (-1.04, 0.51) | -0.65 (-1.24, -0.06) | -1.14 (-1.75, -0.54) | -0.90 (-1.75, -0.06) | -1.38 (-1.82, -0.94) | -1.61 (-2.05, -1.16) | -1.73 (-2.68, -0.78) | -1.89 (-2.50, -1.29) | -3.69 (-4.34, -3.03) |
| -2.11 (-3.16, -1.05) | -1.41 (-2.47, -0.36) | -1.17 (-2.18, -0.17) | -1.09 (-2.15, -0.03) | -0.55 (-1.81,0.70) | -0.27 (-1.20, 0.66) | Carteolol | -0.38 (-1.36, 0.60) | -0.88 (-1.85, 0.08) | -0.64 (-1.79, 0.51) | -1.11 (-2.02, -0.21) | -1.34 (-2.25, -0.43) | -1.47 (-2.69, -0.24) | -1.63 (-2.62, -0.64) | -3.43 (-4.47, -2.39) |
| -2.49 (-3.30, -1.67) | -1.79 (-2.59, -0.99) | -1.55 (-2.29, -0.81) | -1.47 (-2.37, -0.56) | -0.93 (-2.02,0.15) | -0.65 (-1.34, 0.04) | -0.38 (-1.54,0.78) | Brinzolamide | -0.48 (-1.10, 0.14) | -0.25 (-1.26, 0.75) | -0.73 (-1.30, -0.16) | -0.96 (-1.63, -0.29) | -1.08 (-2.20, 0.03) | -1.25 (-2.07, -0.43) | -3.03 (-3.80, -2.26) |
| -2.96 (-3.63, -2.30) | -2.27 (-2.93, -1.61) | -2.03 (-2.57, -1.49) | -1.95 (-2.72, -1.17) | -1.41 (-2.39, -0.43) | -1.13 (-1.63, -0.62) | -0.86 (-1.92,0.20) | -0.48 (-1.17,0.21) | Brimonidine | 0.22 (-0.79, 1.24) | -0.26 (-0.92, 0.41) | -0.48 (-1.16, 0.19) | -0.61 (-1.73, 0.52) | -0.77 (-1.59, 0.05) | -2.55 (-3.36, -1.75) |
| -3.02 (-4.18, -1.86) | -2.33 (-3.49, -1.17) | -2.09 (-3.20, -0.97) | -2.00 (-3.21, -0.80) | -1.47 (-2.81, -0.12) | -1.18 (-2.24, -0.13) | -0.91 (-2.32,0.49) | -0.53 (-1.78,0.72) | -0.06 (-1.22,1.10) | Levobetaxolol | -0.48 (-1.41, 0.44) | -0.71 (-1.63, 0.21) | -0.83 (-2.10, 0.44) | -0.99 (-2.02, 0.03) | -2.77 (-3.72, -1.81) |
| -3.22 (-3.96, -2.48) | -2.53 (-3.27, -1.79) | -2.29 (-2.94, -1.64) | -2.21 (-3.03, -1.39) | -1.67 (-2.70, -0.65) | -1.39 (-1.97, -0.81) | -1.12 (-2.22, -0.02) | -0.74 (-1.51,0.04) | -0.26 (-0.98,0.46) | -0.20 (-1.39, 0.99) | Dorzolamide | -0.23 (-0.73, 0.27) | -0.35 (-1.40, 0.69) | -0.52 (-1.24, 0.21) | -2.31 (-2.99, -1.62) |
| -3.34 (-4.08, -2.60) | -2.65 (-3.39, -1.90) | -2.41 (-3.06, -1.75) | -2.32 (-3.11, -1.53) | -1.79 (-2.81, -0.76) | -1.50 (-2.09, -0.92) | -1.23 (-2.33, -0.14) | -0.85 (-1.71,0.00) | -0.38 (-1.09,0.34) | -0.32 (-1.49, 0.85) | -0.11 (-0.85, 0.62) | Betaxolol | -0.13 (-1.17, 0.92) | -0.29 (-1.03, 0.45) | -2.09 (-2.77, -1.41) |
| -3.54 (-5.29, -1.78) | -2.85 (-4.60, -1.09) | -2.61 (-4.33, -0.88) | -2.52 (-4.31, -0.73) | -1.99 (-3.87, -0.10) | -1.70 (-3.39, -0.02) | -1.43 (-3.36,0.49) | -1.05 (-2.87,0.77) | -0.58 (-2.33,1.18) | -0.52 (-2.50, 1.47) | -0.31 (-2.10, 1.47) | -0.20 (-1.98, 1.58) | Apraclonidine | -0.16 (-1.29, 0.96) | -1.93 (-3.06, -0.81) |
| -3.75 (-4.54, -2.95) | -3.05 (-3.85, -2.26) | -2.81 (-3.48, -2.14) | -2.73 (-3.62, -1.83) | -2.19 (-3.27, -1.12) | -1.91 (-2.58, -1.24) | -1.64 (-2.79, -0.49) | -1.26 (-2.20, -0.32) | -0.78 (-1.59,0.02) | -0.73 (-1.97, 0.52) | -0.52 (-1.39, 0.34) | -0.41 (-1.27, 0.46) | -0.21 (-2.02, 1.61) | Unoprostone | -1.79 (-2.65, -0.94) |
| -5.60 (-6.31, -4.88) | -4.90 (-5.64, -4.17) | -4.66 (-5.32, -4.01) | -4.58 (-5.35, -3.81) | -4.04 (-5.07, -3.02) | -3.76 (-4.34, -3.18) | -3.49 (-4.58, -2.40) | -3.11 (-3.94, -2.28) | -2.63 (-3.35, -1.92) | -2.58 (-3.74, -1.41) | -2.37 (-3.09, -1.66) | -2.26 (-2.96, -1.55) | -2.06 (-3.84, -0.28) | -1.85 (-2.68, -1.02) | Placebo |

Figure S1: Network meta-analysis for the IOP data. Results are the mean difference in the column-defining treatment compared with the mean difference in the row-defining treatment. The lower triangular matrix refers to the standard NMA based on Lu and Ades' approach, and upper triangular matrix refers to the proposed method.

| Bimatoprost | Travoprost | Latanoprost | Levobunolol | Tafluprost | Timolol | Carteolol | Brinzolamide | Brimonidine | Levobetaxolol | Dorzolamide | Betaxolol | Apraclonidine | Unoprostone | Placebo |
|---|---|---|---|---|---|---|---|---|---|---|---|---|---|---|
| Bimatoprost | -0.72 (-1.12, -0.31) | -0.94 (-1.35, -0.53) | -1.03 (-1.93, -0.13) | -1.59 (-2.56, -0.61) | -1.85 (-2.23, -1.47) | -2.11 (-2.97, -1.25) | -2.49 (-3.16, -1.82) | -2.98 (-3.66, -2.30) | -2.75 (-3.66, -1.83) | -3.22 (-3.77, -2.67) | -3.45 (-4.02, -2.89) | -3.58 (-4.59, -2.56) | -3.74 (-4.40, -3.08) | -5.53 (-6.24, -4.82) |
| -0.53 (-1.15, 0.08) | Travoprost | -0.20 (-0.55, 0.15) | -0.32 (-1.19, 0.56) | -0.87 (-1.83, 0.08) | -1.12 (-1.44, -0.79) | -1.39 (-2.23, -0.55) | -1.77 (-2.41, -1.14) | -2.27 (-2.91, -1.63) | -2.03 (-2.92, -1.13) | -2.50 (-3.02, -1.99) | -2.73 (-3.27, -2.20) | -2.86 (-3.86, -1.86) | -3.02 (-3.65, -2.39) | -4.82 (-5.51, -4.12) |
| -0.92 (-1.69, -0.14) | -0.11 (-0.49, 0.27) | Latanoprost | -0.12 (-0.99, 0.76) | -0.67 (-1.62, 0.28) | -0.92 (-1.26, -0.57) | -1.19 (-2.03, -0.35) | -1.57 (-2.21, -0.93) | -2.07 (-2.71, -1.43) | -1.83 (-2.72, -0.93) | -2.30 (-2.81, -1.79) | -2.53 (-3.07, -2.00) | -2.66 (-3.66, -1.65) | -2.82 (-3.38, -2.25) | -4.61 (-5.31, -3.92) |
| | | | Levobunolol | -0.56 (-1.77, 0.65) | -0.90 (-1.69, -0.10) | -1.08 (-2.19, 0.02) | -1.47 (-2.48, -0.46) | -1.95 (-2.96, -0.93) | -1.72 (-2.90, -0.54) | -2.20 (-3.15, -1.26) | -2.43 (-3.36, -1.51) | -2.55 (-3.79, -1.31) | -2.72 (-3.74, -1.69) | -4.50 (-5.68, -3.32) |
| | | | | Tafluprost | -0.40 (-1.35, 0.54) | -0.52 (-1.72, 0.67) | -0.91 (-1.99, 0.17) | -1.37 (-2.45, -0.29) | -1.16 (-2.40, 0.08) | -1.65 (-2.65, -0.64) | -1.87 (-2.89, -0.86) | -1.99 (-3.31, -0.68) | -2.16 (-3.24, -1.07) | -3.93 (-5.04, -2.81) |
| -2.09 (-2.48, -1.71) | -0.93 (-1.30, -0.57) | -1.26 (-1.68, -0.85) | 0.03 (-0.39, 0.44) | -0.16 (-1.12, 0.80) | Timolol | -0.26 (-1.04, 0.51) | -0.65 (-1.24, -0.06) | -1.14 (-1.75, -0.54) | -0.90 (-1.75, -0.06) | -1.38 (-1.82, -0.94) | -1.61 (-2.05, -1.16) | -1.73 (-2.68, -0.78) | -1.89 (-2.50, -1.29) | -3.69 (-4.34, -3.03) |
| | | | -2.90 (-4.59, -1.22) | | 0.03 (-0.61, 0.68) | Carteolol | -0.38 (-1.36, 0.60) | -0.88 (-1.85, 0.08) | -0.64 (-1.79, 0.51) | -1.11 (-2.02, -0.21) | -1.34 (-2.25, -0.43) | -1.47 (-2.69, -0.24) | -1.63 (-2.62, -0.64) | -3.43 (-4.47, -2.39) |
| | -2.70 (-3.99, -1.41) | | | | -0.78 (-2.61, 1.04) | | Brinzolamide | -0.48 (-1.10, 0.14) | -0.25 (-1.26, 0.75) | -0.73 (-1.30, -0.16) | -0.96 (-1.63, -0.29) | -1.08 (-2.20, 0.03) | -1.25 (-2.07, -0.43) | -3.03 (-3.80, -2.26) |
| | -1.20 (-3.77, 1.37) | -1.08 (-2.12, -0.05) | | | -1.33 (-2.37, -0.28) | | -1.09 (-1.92, -0.26) | Brimonidine | 0.22 (-0.79, 1.24) | -0.26 (-0.92, 0.41) | -0.48 (-1.16, 0.19) | -0.61 (-1.73, 0.52) | -0.77 (-1.59, 0.05) | -2.55 (-3.36, -1.75) |
| | | | | | -1.25 (-2.23, -0.27) | | | | Levobetaxolol | -0.48 (-1.41, 0.44) | -0.71 (-1.63, 0.21) | -0.83 (-2.10, 0.44) | -0.99 (-2.02, 0.03) | -2.77 (-3.72, -1.81) |
| | | -2.90 (-3.70, -2.10) | | | -1.20 (-1.88, -0.52) | | -0.32 (-0.80, 0.17) | | | Dorzolamide | -0.23 (-0.73, 0.27) | -0.35 (-1.40, 0.69) | -0.52 (-1.24, 0.21) | -2.31 (-2.99, -1.62) |
| | | -1.05 (-2.62, 0.51) | -4.73 (-10.01, 0.55) | | -1.70 (-2.41, -0.99) | | | 0.04 (-0.95, 1.03) | -2.00 (-3.54, -0.46) | -0.30 (-0.96, 0.36) | Betaxolol | -0.13 (-1.17, 0.92) | -0.29 (-1.03, 0.45) | -2.09 (-2.77, -1.41) |
| | | | | | -1.76 (-3.27, -0.26) | | | | | | | Apraclonidine | -0.16 (-1.29, 0.96) | -1.93 (-3.06, -0.81) |
| | | | | | -1.43 (-2.47, -0.39) | | | | | | | | Unoprostone | -1.79 (-2.65, -0.94) |
| -4.60 (-5.60, -3.60) | | | -7.51 (-8.53, -6.50) | | -3.61 (-4.63, -2.59) | | -2.28 (-4.04, -0.52) | -2.30 (-3.99, -0.61) | -3.00 (-4.53, -1.47) | -1.33 (-1.68, -0.98) | -2.28 (-3.65, -0.91) | | -0.50 (-1.70, 0.70) | Placebo |

Figure S2: Network meta-analysis for the IOP data. Results are the mean difference in the column-defining treatment compared with the mean difference in the row-defining treatment. The lower triangular matrix refers to the pairwise meta-analysis method, and the upper triangular matrix refers to the proposed method.

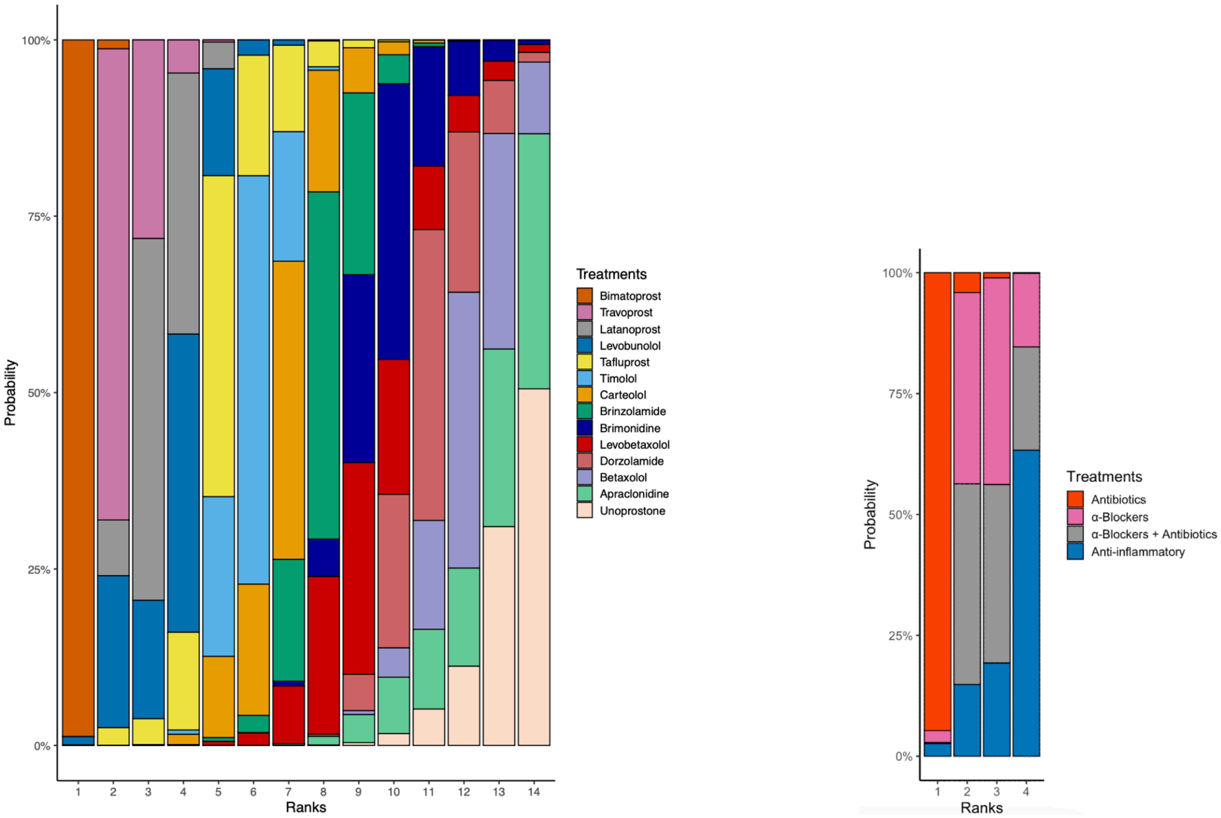


(a) Adapted from IOP data (b) Adapted from the CP/CPPS data

Figure S3: Treatment ranking was calculated based on the surface under the cumulative ranking (SUCRA) method.

| Antibiotics | α-blockers | α-blockers + Antibiotics | Anti-inflammatory | Placebo |
|---|---|---|---|---|
| Antibiotics | -3.25 (-6.82, 0.32) | -3.44 (-6.08, -0.80) | -4.13 (-8.89, 0.63) | -7.75 (-12.40, -3.10) |
| -3.07 (-7.32, 1.17) | α-blockers | -0.25 (-2.98, 2.47) | -0.97 (-4.41, 2.47) | -4.34 (-7.67, -1.00) |
| -3.26 (-8.44, 1.92) | -0.19 (-5.25, 4.88) | α-blockers + Antibiotics | -0.69 (-4.57, 3.18) | -4.28 (-8.08, -0.49) |
| -4.68 (-9.90, 0.53) | -1.61 (-6.33, 3.10) | -1.42 (-7.69, 4.84) | Anti-inflammatory | -3.11 (-4.48, -1.74) |
| -7.84 (-11.78, -3.89) | -4.76 (-8.01, -1.51) | -4.58 (-9.82, 0.67) | -3.15 (-6.57, 0.26) | Placebo |

Figure S4: Network meta-analysis for the CP/CPPS data. Results are the mean difference in the column-defining treatment compared with the mean difference in the row-defining treatment. The lower triangular matrix refers to the standard NMA using the Lu and Ades' approach, and the upper triangular matrix refers to the proposed method.
